# Differential transcript usage unravels gene expression alterations in Alzheimer’s disease human brains

**DOI:** 10.1101/2020.03.19.20038703

**Authors:** Diego Marques-Coelho, Lukas Iohan da Cruz Carvalho, Ana Raquel Melo de Farias, NeuroCEB Brain Bank, Jean-Charles Lambert, Marcos Romualdo Costa

## Abstract

Alzheimer’s disease (AD) is the leading cause of dementia in aging individuals. However pathophysiological processes involved in the brain are still poorly understood. Among numerous strategies, a comprehensive overview of gene expression alterations in the diseased brain has been proposed to help for a better understanding of the disease processes. In this work, we probed the differential expression of genes in different brain regions of healthy and AD adult subjects using data from three large studies: Mayo Clinic; Mount Sinai Brain Bank (MSBB) and ROSMAP. Using a combination of differential expression of gene and isoform switch analyses we provide a detailed landscape of gene expression alterations in the temporal and frontal lobes, harboring brain areas affected at early and late stages of the AD pathology, respectively. Next, we took advantage of an indirect approach to assign the complex gene expression changes revealed in bulk RNAseq to individual cell types of the adult brain. This strategy allowed us to identify cell type/subtype specific isoform switches in AD brains previously overlooked. Among these alterations, we show isoform switches in the AD causal gene APP (Amyloid Beta Precursor Protein) and the risk gene BIN1 (Bridging Integrator 1), which could have important functional consequences in neuronal cells. Altogether, our work proposes a novel integrative strategy to analyze RNAseq data in AD based on both gene/transcript expression and regional/cell-type specificities.

## Introduction

Changes in gene expression characterize a multitude of human diseases and have been successfully used to predict molecular and cellular mechanisms associated with pathological processes (Masters et al., 2015). Alzheimer’s disease (AD) is the most prevalent type of dementia and causes a progressive cognitive decline, for which there is no effective treatment or cure. Although expression analyses in brain diseases are generally limited by tissue availability, RNA sequencing (RNAseq) data have been generated from *postmortem* brain samples of healthy and AD individuals (Allen et al., 2016; De Jager et al., 2018; Wang et al., 2018). However, a comprehensive description of the gene expression alterations in the AD brain remains elusive.

Recent work has begun to address this important gap in the study of AD pathology using bulk brain tissue RNA sequencing (RNAseq) (Raj et al., 2018) or single-cells RNA sequencing (scRNAseq) (Grubman et al., 2019; Mathys et al., 2019). However, these studies have focused on samples obtained from different brain regions, namely the dorsolateral prefrontal (Mathys et al., 2019; Raj et al., 2018) and entorhinal cortices (Grubman et al., 2019), which could lead to important discrepancies in the results. In fact, AD pathology shows a progressive impact on different brain regions, characterized at early stages by the presence of TAU protein inclusions in the locus coeruleus, the transentorhinal and entorhinal regions (stages I and II). This is followed by the presence of TAU inclusions in the hippocampal formation and some parts of the neocortex (stages III and IV), followed by large parts of the neocortex (stages V and VI) (Braak & Braak, 1991). This temporal progression of AD pathology could differently impact gene expression in those brain areas. Accordingly, a recent study has shown that changes in protein expression are much more prominent in areas affected at early and intermediate stages, such as the hippocampus, entorhinal cortex and cingulate cortex in the temporal lobe, compared to other brain regions affected at later stages of AD pathology, such as sensory cortex, motor cortex and cerebellum (Xu et al., 2019).

Another important aspect to consider is the descriptive relevance of gene expression analysis based solely on the identification of differentially expressed genes (DEG), which fails to detect dynamics in the expression of multiple related transcripts (Yi et al., 2018). Recently, new approaches using transcripts-level analysis, so called differential transcript usage (DTU), enables identification of alternative splicing and isoform switches with prediction of functional consequences (Anders et al., 2012; Vitting-Seerup & Sandelin, 2019). Therefore, important gene expression modifications in the AD brain could occur at the transcript level and be overlooked in classical DEG analyses.

Here, we took advantage of three available RNAseq datasets, generated using samples from different brain regions, to systematically probe gene expression changes (DEG and DTU) in AD. In the Mayo’s clinic study, both the temporal cortex and cerebellum were used to obtain bulk RNAseq (Allen et al., 2016). In the Religious Orders Study (ROS) and Memory and Aging Project (MAP), henceforth called ROSMAP, the dorsolateral prefrontal cortex was used (De Jager et al., 2018). Finally, in the Mount Sinai/JJ Peters VA Medical Center Brain Bank (MSBB), 4 different Brodmann areas of the brain were studied: areas 22 and 36 from the temporal lobe, areas 10 and 44 in the frontal lobe (Wang et al., 2018). We also added another level of complexity using an indirect approach to assign DEGs and gDTUs to unique cell types in order to identify AD gene expression signatures for neural cells, microglia and endothelial cells. Finally, we linked these alterations with AD causal and risk genes, identifying novel isoform switches in BIN1 and APP genes of potential functional consequences for pathology progression.

## Results

### Regional gene expression alterations in the AD brain correlates with pathological progression

Several consortia have generated RNAseq data from brains of individuals with a clinical and/or pathological diagnostic of AD (Allen et al, 2016; Wang et al, 2018; De Jager et al, 2018). Considering the regional progression of AD pathology (Braak and Braak, 1991), we set out to identify and compare differentially expressed genes (DEG) in the temporal lobe (TL), encompassing brain regions affect at early stages of the AD such as the hippocampus and entorhinal cortex, and in the frontal lobe (FL), affect at more advanced stages of the pathology (Figure 1). Comparisons between control and AD individuals were performed independently for each dataset and only genes with fold change > 1.3 and FDR > 0.01 were considered as DEGs. We found 3,348 (1244 down- and 2104 up-regulated genes) and 2,172 (1170 down and 999 up-regulated genes in BM22 and BM36; 3 genes regulated in opposite directions in these two areas) DEGs in the TL of AD individuals compared to their respective controls in the MSBB_TL and Mayo datasets, respectively (Figure 2A-B; Supplementary table 1). Of those DEGs, 734 genes (145 down and 520 up) were commonly regulated in both Mayo and MSBB_TL (88.4% of genes altered in the same direction; 15,33% of overlap; p= 8.56 × 10^−59^, hypergeometric test). In contrast, only 327 (113 down and 214 up) and 209 (97 down and 112 up) DEGs were detected in the MSBB_FL and ROSMAP, respectively. Of those, 31 genes (18 down and 13 up) were found in both datasets (7,34% of overlap; p = 1.67 × 10^−14^, hypergeometric test) (Figure 2A-B; Supplementary table 1). This small number of DEGs in the FL is in agreement with previous data obtained from the DLPFC (106 down- and 158 up-regulated genes with FC>1.3; Canchi et al., 2019). Among DEGs detected in the FL, 62.5% were also detected in the TL (Figure 2B), suggesting that similar molecular changes occur in these brain areas, but at different stages of the disease progression. The differences in the number of DEGs detected in the FL and TL can neither be attributed to lack of statistical power nor potential biases due to tissue processing, since the number of samples in the FL is larger than in the TL groups (Figure 1) and differences are observed even in samples obtained from the same donors (compare MSBB_TL and MSBB_FL in Figure 2). Thus, changes in gene expression are much more prominent in brain areas affected at early stages of AD pathology.

**Figure 1.**
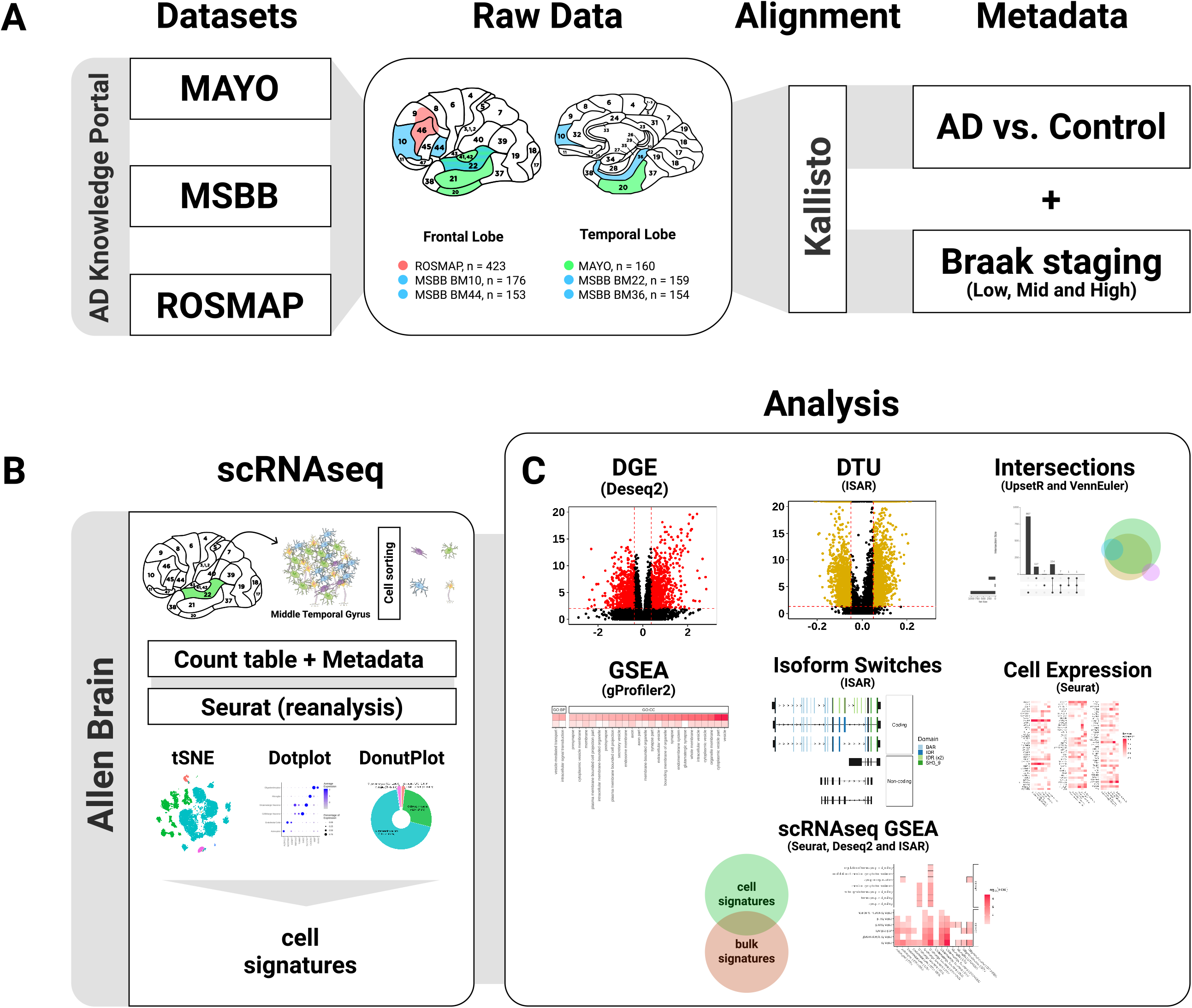
Schematic summary of the methodology. Datasets obtained from 3 consortia (Mayo, MSBB and ROSMAP) were grouped according to the brain region sampled in frontal lobe (FL) or temporal lobe (TL). Next, RNAseq data was pseudo-aligned using Kallisto and analyzed using the packages from R (version 3.6) DESeq2, IsoformSwitchAnalyzeR (ISAR) and gene-set enrichment analysis (GSEA). Assignment of differentially expressed genes or isoform switches to specific cell types was performed indirectly using scRNAseq signatures obtained from the Allen Brain Atlas transcriptomic data.

**Figure 2.**
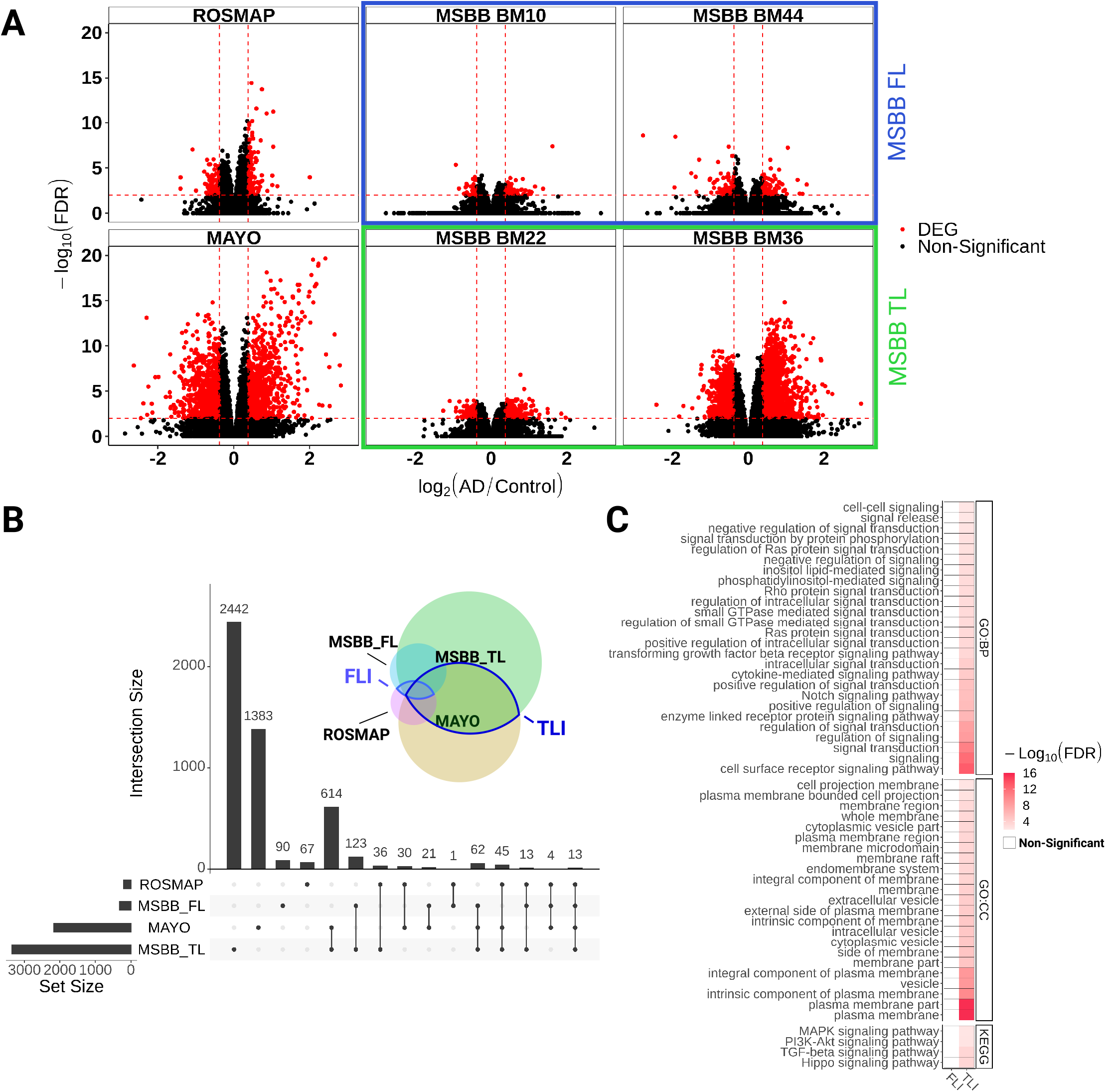
Gene expression alterations are more prominent in the temporal than frontal lobe of AD patients. A) Volcano plots showing differentially expressed genes (DEG, red dots; FC > 1.3 and FDR < 0.01) in the frontal lobe (ROSMAP and MSBB FL - BM10 and BM44) and temporal lobe (Mayo and MSBB TL - BM22 and BM36). B) Upset plot showing the total number of DEGs identified in each dataset (horizontal bars) and the number of DEGs exclusive of one dataset (first four vertical bars) or shared by different datasets (other vertical bars). Black dots below vertical bars indicate datasets quantified. Venn diagram illustrates the same results in colors and circle sizes. C) Gene ontology terms enriched for DEGs identified in the TL or FL intersections (TLI and FLI, respectively).

To select genes consistently altered in AD brains, considering the several sources of measurement variations in RNAseq experiments (Van den Berge et al., 2017), we decided to focus only on DEGs replicated in at least two independent datasets obtained from related brain areas. This resulted in a set of 734 DEGs detected in both Mayo and MSBB TL (temporal lobe intersection - TLI), and 31 DEGs shared between ROSMAP and MSBB FL (frontal lobe intersection - FLI) (Supplementary table 2). Among TLI DEGs we observed ABCA1 and 2 (ATP Binding Cassette Subfamily A Member 1 and 2), primarily involved in the maintenance of normal brain homeostasis and associated with AD and other neurological diseases (Abuznait & Kaddoumi, 2012); Complement C1R and C1S, involved in the immune/inflammatory response and previously shown to be upregulated in the brain of a 3 × Tg mouse model of AD when Aβ plaques start to accumulate (Benoit et al., 2013); REST (RE1 Silencing Transcription Factor), which regulates neural circuit activity during aging (Zullo et al., 2019); GAD1 and 2 (Glutamate Decarboxylase 1 and 2), SLC32A1 (Solute Carrier Family 32 GABA Vesicular Transporter, Member 1), CALB1 (Calbindin 1), PVALB (Parvalbumin), SST (Somatostatin) and VIP (Vasoactive Intestinal Peptide), all expressed in GABAergic neurons and involved in cognitive decline in AD and other neurological diseases (Prévot & Sibille, 2020). Among the few DEGs common to TLI and FLI, we observed a significant downregulation of the neurosecretory protein VGF (VGF Nerve Growth Factor Inducible), recently suggested as a key regulator of Alzheimer’s disease (Beckmann et al., 2020).

Next, we used gene set enrichment analyses (GSEA) to assess the functional profile of the DEGs identified in our analysis. Again, we used only genes commonly altered in two datasets (TLI or FLI) to avoid inaccurate results associated with the use of large gene sets in functional analysis (Subramanian et al., 2005). We found that TLI DEGs were significantly enriched for terms (GO:BP, GO:CC and KEGG) associated with generic biological processes, such as cell-signaling pathways and cell-cell signaling, whereas the small number of DEGs in the FLI were not significantly enriched for any term (Figure 2C; Supplementary table 3). The limited number of significant gene set enrichment observed in our analysis after inputting DEGs is in disagreement with results reported by Canchi et al. (2019). This discrepancy can likely be explained by the use of stringent criteria to detect TLI DEGs in our study (only genes detected in at least two independent datasets with FC>1.3 and FDR<0.01), which significantly reduce the number of genes used in the GSEA.

### Differential transcript usage analysis reveals novel genes associated with AD pathology

Gene-level expression analysis lacks the sensitivity to detect possible changes at the transcript-level caused, for example, by alterations in alternative splicing (Vitting-Seerup & Sandelin, 2017; Yi et al., 2018). To overcome this limitation, we used differential transcript usage (DTU) analysis to identify additional alterations of gene expression in the AD brains compared to controls. We observed 2,509 and 1,843 genes with differential transcript usage (gDTU) in the temporal lobe of AD brains studied in the Mayo and MSBB datasets, respectively (Figure 3A-B; Supplementary table 1). Similar to what we observed for DEGs, a much smaller number of gDTUs were detected in the frontal lobe, both in ROSMAP and MSBB studies (59 and 855 genes with transcripts altered, respectively). We found 435 gDTUS in TLI (11,1% of overlap; p= 6.16 × 10^−25^, hypergeometric test) and 13 gDTUs in FLI (1,47% of overlap; p= 2.56 × 10^−3^, hypergeometric test) (Supplementary table 2). In TLI, most gDTUs did not overlap with DEGs (TL - 34 gDTUs that are DEGs out of 435 gDTUs, Figure 4A), whereas in FLI, we found no overlap at all. Consistent with this small overlap, GSEA using only DEGs, only gDTUs or both showed complementary results (Figure 4B). GSEA using gDTUs (alone or in combination with DEGs) showed significant enrichment for vesicle-mediated transport and other synapse-related terms, which were not observed while inputting only DEGs (Figure 3C and 4B; Supplementary table 3). The functional enrichment annotation using both DEGs and gDTUs is in agreement with previous studies using scRNAseq to identify gene expression alterations in unique cell types (Mathys et al., 2019; Grubman et al., 2019) and clearly improves the annotation observed using only DEGs, suggesting that the use of DTU analysis could contribute to unraveling gene expression alterations overlooked in the classical DEG analysis.

**Figure 3.**
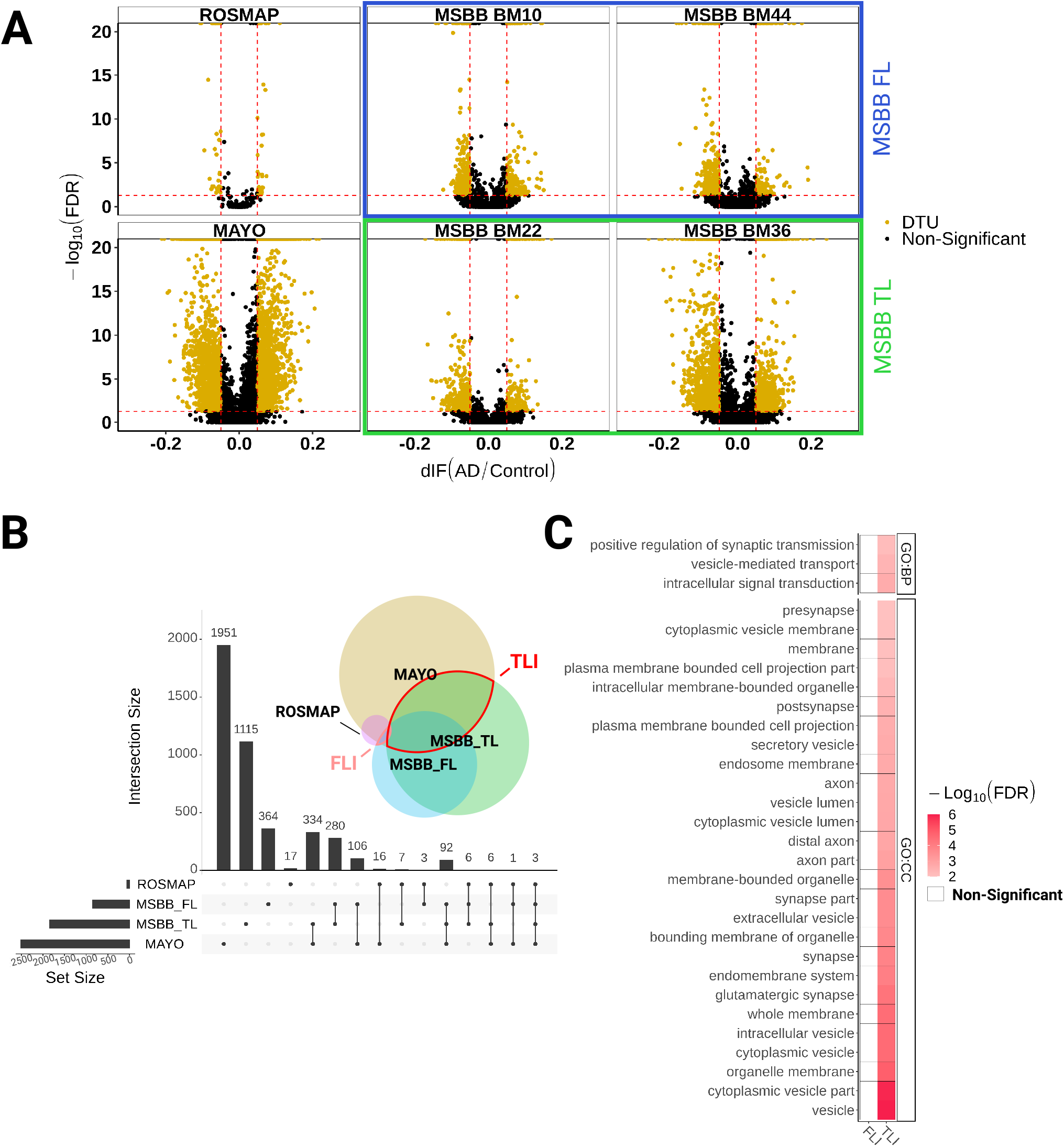
Differential transcript usage analysis identifies gene expression alterations in AD associated with synapse transmission. A) Volcano plots showing genes with differential transcript usage (gDTU, yellow dots; Differential isoform fraction (dIF) > 0.05 and FDR < 0.05) in the frontal lobe (ROSMAP and MSBB_FL - BM10 and BM44) and temporal lobe (Mayo and MSBB_TL - BM22 and BM36). B) Upset plot showing the total number of gDTUs identified in each dataset (horizontal bars) and the number of gDTUs exclusive of one dataset (first four vertical bars) or shared by different datasets (other vertical bars). Black dots below vertical bars indicate datasets quantified. Venn diagram illustrates the same results in colors and circle sizes.C) Synapse-related terms enriched for gDTUs in the TLI are not observed in the FLI.

**Figure 4.**
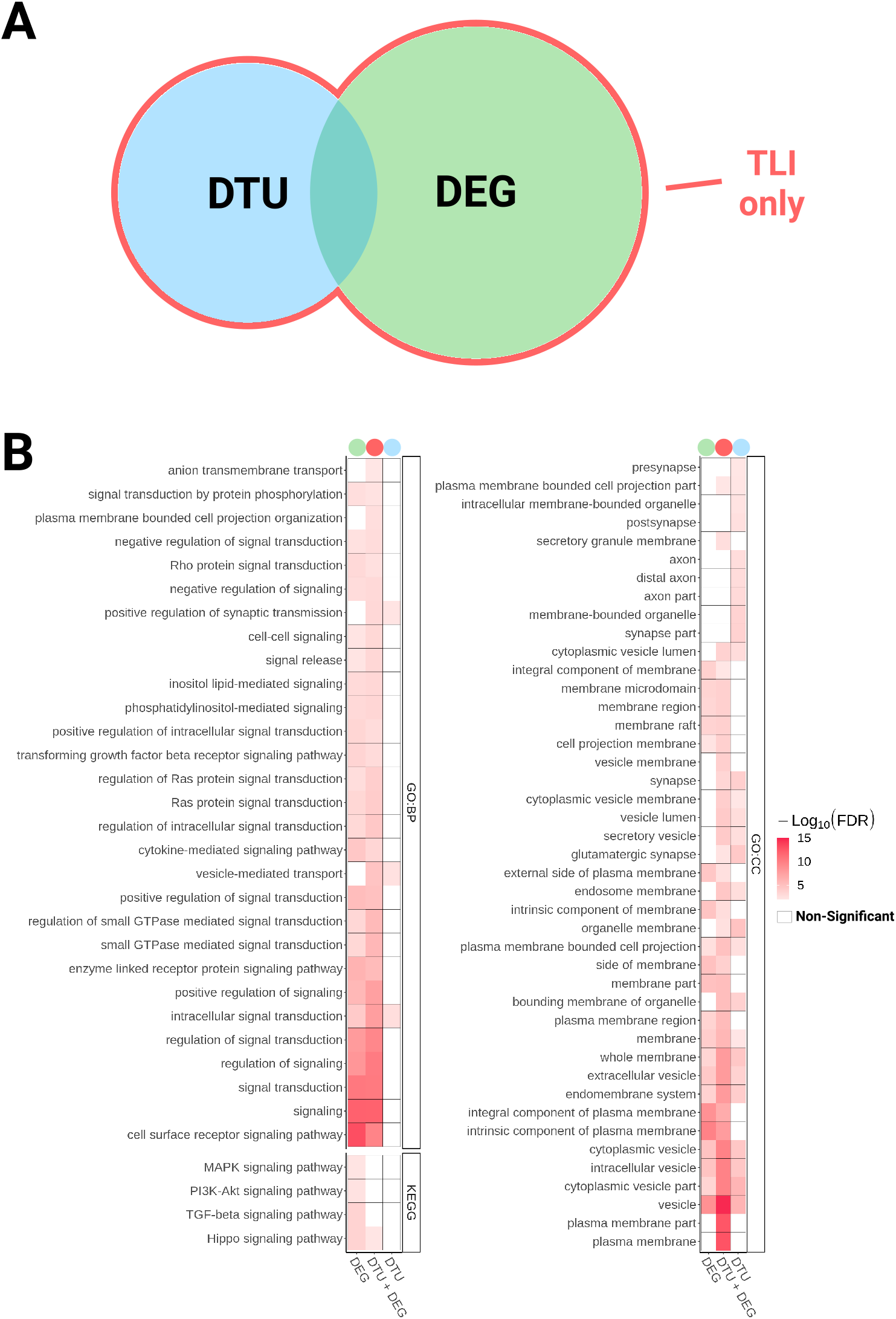
Differential transcript usage analysis in AD brains reveals gene expression alterations overlooked in DEG analysis. A) Venn diagram showing DEGs and gDTUs identified in the TLI. B) Comparison of GO and KEGG terms enriched for DEG, gDTU or DEG+gDTU identified in the TLI.

Among genes with isoform switches enriched in synaptic-related terms, we observed the AD causal gene APP, previously associated with regulation of synapse transmission and long-term plasticity in AD (Kamenetz et al., 2003); NSG1 (Neuronal Vesicle Trafficking Associated 1), which has been implicated in the regulation of AMPA receptors (AMPAR) and APP trafficking, thus affecting synaptic transmission, plasticity and Aβ production (Alberi et al., 2005; Norstrom et al., 2010); RELN (Reelin) that plays important role in synaptic transmission and has been associated with AD (Yu et al., 2016); GABRA1 (Gamma-Aminobutyric Acid Type A Receptor Subunit Alpha1), which encodes for a subunit of the main ionotropic GABA receptor in the brain and has previously been shown to be downregulated in the AD brain (Limon et al., 2012).

### Alternative splice events in AD brains and functional consequences

To identify the causes subjacent to gene isoform switches in the AD brain, we quantified the frequency of splicing events associated with the isoform switches detected in AD compared to control brains (Figure 5; Supplementary table 4). We found that alternative transcription start site (ATSS), alternative transcription termination site (ATTS) and exon skipping (ES) were the most frequent splicing events in AD brains (Figure 5B). Other common splicing events observed were alternative 3’ or 5’ splice sites (A3 and A5, respectively), multiple exon skipping (MES) and intron retention (IR) (Figure 5B). These observations suggest that changes in alternative splicing could be implicated in AD pathogenesis, corroborating previous analyses in the ROSMAP cohort using intronic usage ratios to identify abnormal splicing events in the AD brain (Raj et al., 2018).

**Figure 5.**
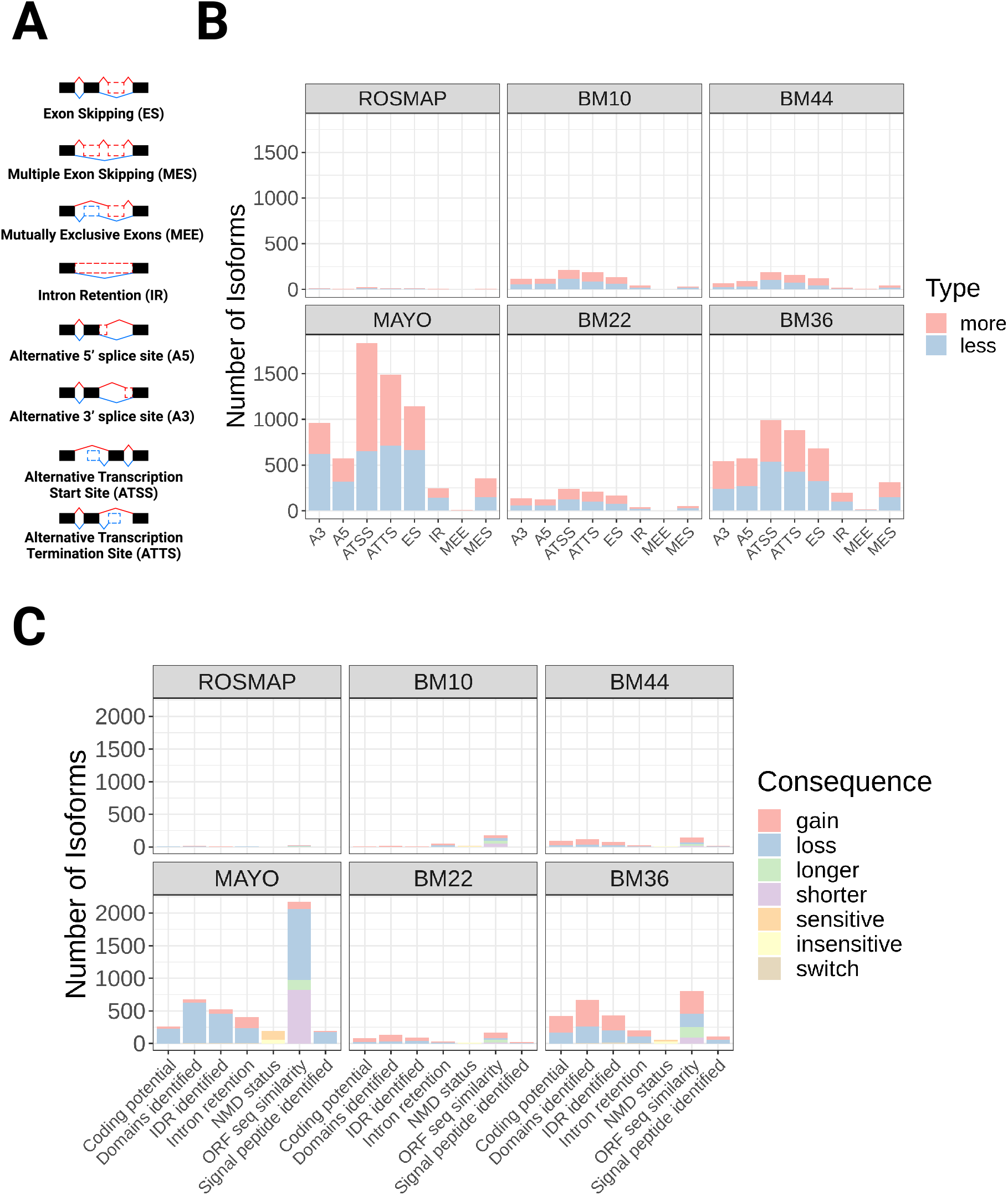
Alternative splicing mechanisms associated with isoform switches and consequences for protein expression. A) Schematic showing different splicing events that can lead to gene isoform switches. B) Quantification of the number of isoforms showing more or less splicing events in AD compared to controls for each dataset. C) Quantification of the number of isoforms showing i) gain or loss of coding potential, domains/signal peptides identified, intrinsically disordered regions (IDR), intron retention, open reading frame (ORF) sequencing similarity; ii) switch (simultaneous gain and loss) of domains identified or IDR; iii) sensitive or insensitive to nonsense mediated decay (NMD); and iv) longer or shorter ORF sequencing similarity.

Alternative splicing events may have diverse functional consequences for protein expression, such as shifting the frequency of transcripts containing introns (non-coding) or mRNA stability (nonsense mediated decay) or leading to gain/loss of protein domains, intrinsically disordered regions or signaling peptides (Vitting-Seerup & Sandelin, 2019). Quantification of these consequences revealed some interesting differences between Mayo and MSBB BM36 (Figure 5C), the two datasets with largest numbers of gDTUs. Whereas in the Mayo dataset, a high number of isoforms showed loss of coding potential and protein domains, in the MSBB BM36 isoforms showed an even distribution of loss and gain of coding potential or protein domains (Figura 5B). These differences could be at least partly explained by the larger number of gDTUs detected in the Mayo compared to MSBB TL (Figure 3) and are likely related to the different median read depth of these datasets (Mayo - 12.58 billion bases; MSBB BM22 - 3.23 billion bases; MSBB BM36 - 3.56 billion bases; Wan et al., 2020).

### Differential expression of genes involved in alternative splicing correlates with isoform switches during disease progression

To evaluate whether the emergence of gDTUs could be correlated with AD pathology hallmarks, we quantified the total of gDTUs observed at different disease stages in the MSBB dataset using the Braak classification (Figure 6; Supplementary Table 5). For this purpose, we subdivided samples in three groups: low Braak (0, 1 and 2) - 196 samples (clinical diagnosis: 15 AD and 181 controls); mid Braak (3 and 4) - 133 samples (clinical diagnosis: 58 AD and 75 controls); and high Braak (5 and 6) - 308 samples (clinical diagnosis: 305 AD and 3 controls). Next, we evaluated the number of gDTUs when comparing individuals at these different stages (Figure 6). We observed that most gDTUs were detected only while comparing high with either low or mid Braak stages (Figure 6A-D). This pattern was observed both in the FL (BM10 and BM44) and TL (BM22 and BM36), suggesting that gene isoform switches positively correlate with AD pathology progression.

**Figure 6.**
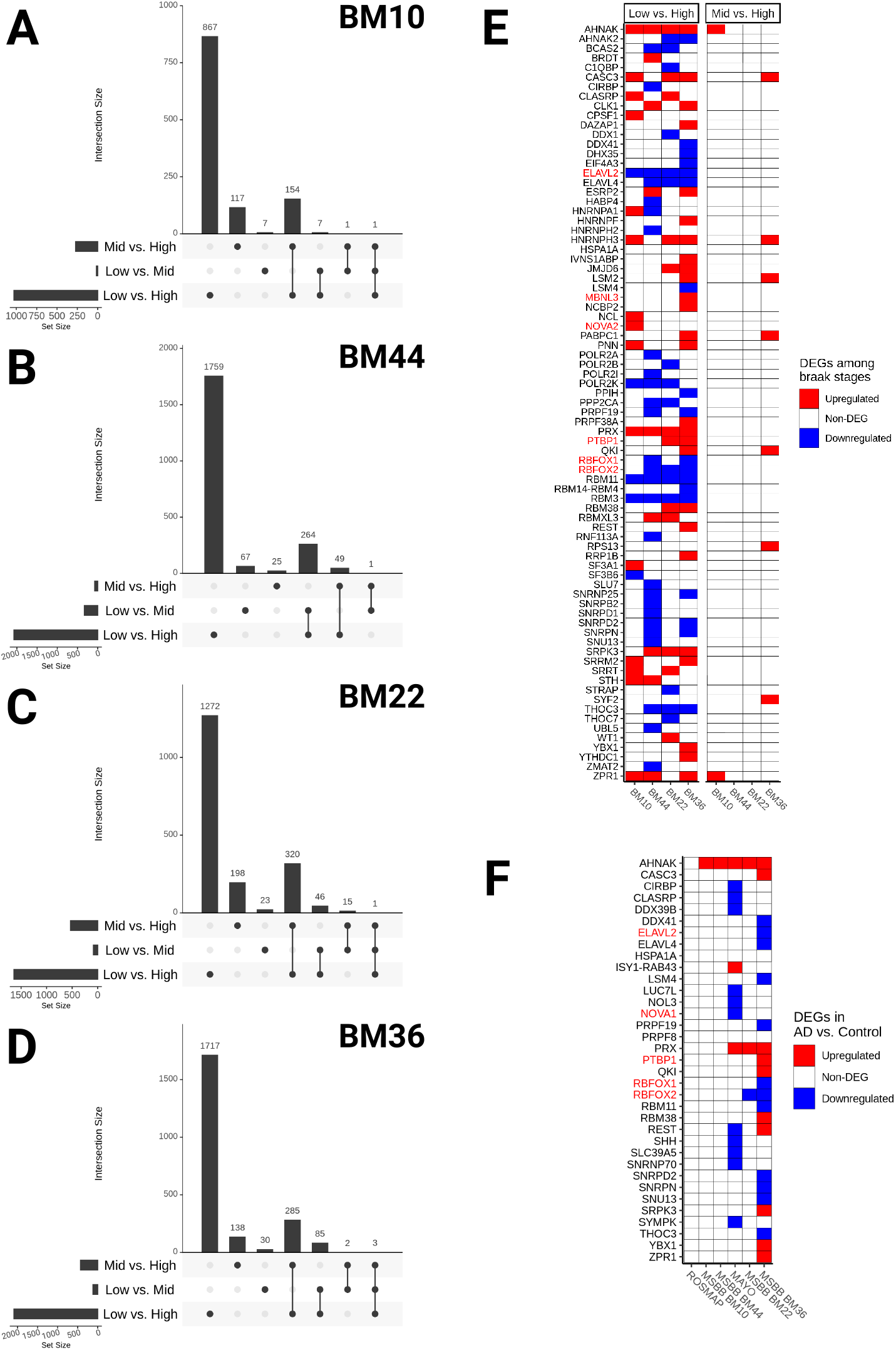
Coincidence between altered expression of splicing-related genes and gDTUs in advanced pathologic stages of AD. A-D) Upset plots showing the total number of gDTU identified in the comparison between different Braak stages (Low vs. High, Low vs. Mid and Mid vs. High) in BM10 (A), BM44 (B), BM22 (C) or BM36 (D). Horizontal bars show the total number of gDTUs identified in each comparison (Low vs. High, Low vs. Mid and Mid vs. High), whereas vertical bars indicate the gDTUs exclusive or common to different comparisons. Black dots below vertical bars indicate stages analyzed E-F) Differential expression of genes associated with splicing/spliceosome after comparison of different Braak stages (E) or AD vs Controls in different datasets (F). Red and blue squares indicate, respectively, up and downregulated genes. Gene symbols highlighted in red indicate genes belonging to the neuronal splicing machinery.

Next, we set out to evaluate alterations in the expression of genes encoding for proteins of the splicing machinery between the same Braak stages. We found that among 441 genes related to ‘splicing’ or ‘spliceosome’ terms (Supplementary table 6), 79 were DEGs at high compared to low or mid Braak stages (Figure 6E). In contrast, we could not detect any DEG in the comparison of mid vs low Braak stages. Among DEGs detected in the comparison between high and low/mid Braak stages, we observed that several genes specifically associated with the neuronal splicing regulatory network (Raj & Blencowe, 2015), such as RBFOX and 2 (RNA Binding Fox-1 Homolog 1 and 2), ELAVL2 (ELAV Like RNA Binding Protein 2), MBNL3 (Muscleblind Like Splicing Regulator 3), PTBP1 (Polypyrimidine Tract Binding Protein 1) and NOVA2 (NOVA Alternative Splicing Regulator 2) (Figure 6E, highlighted in red). A similar correlation between pathological burden and differential expression of the same 441 splice-related genes was observed in the comparison between all AD versus control subjects of the different datasets (Figure 6F). Changes in the expression of those genes were hardly observed in FL (low number of gDTUs – Figure 3), but were frequent in TL samples (high number of gDTUs – Figure 3), albeit to a lesser extent than that observed in the comparison between different Braak stages (likely due to the effects of combining low, mid and high Braak stages in the AD group). Remarkably, the majority of the splicing-related genes with altered expression in the Mayo dataset was not reproduced in the MSBB BM36 dataset, and vice-versa (Figure 6F). This could help to explain the dissimilar consequences of alternative splicing events observed in those datasets (Figure 5C) and suggest that a myriad of proteins could be involved in altered splicing in the AD brains.

### Differential gene expression in separate cell types of the human brain

Considering the cellular diversity in the brain, we took an indirect approach to sort DEGs and gDTUs according to individual cell types. To that, we used scRNAseq data obtained from the adult human brain to identify cell types expressing the genes altered in our DEG/gDTU analysis (Figure 7; Supplementary Figure 1). We found that, out of the 1135 genes with altered expression, i.e. gDTU + DEG, in the TLI (Figures 2 and 3), 839 were found in at least one cell-type using as cut-off the expression in more than 10% of cells assigned for a specific cell-type (Supplementary table 7). From these, 239 were identified in unique cell-types/subtypes, 396 in multiple (2-4 cell-types) and 211 in all cell-types analyzed (Figure 7A; Supplementary Figures 1 and 2; Supplementary table 7). Confirming the efficacy of our strategy, GO analyses using cell-type specific genes revealed that DEGs/gDTUs in the TLI of AD patients were significantly enriched for biological processes associated with inflammation in microglial cells, whereas those associated with cell adhesion were enriched in endothelial cells (Figure 7B; Supplementary table 8). Similarly, DEGs/gDTUs identified in neuronal cells were enriched for GO terms such as synaptic signaling, synaptic plasticity and synapse vesicle cycle (Figure 7C). Notably, these enrichments were more significant in GABAergic neurons, which could suggest a more pronounced pathological burden on these cells compared to glutamatergic neurons (Figure 7C). Comparison of the cell-type gene expression signatures identified in our work with previous studies using scRNAseq in AD (Mathys et al., 2019; Grubman et al., 2019) showed a similar degree of overlap (Supplementary Figures 3 and 4; Supplementary table 9), further supporting the effectiveness of our strategy to assign gene expression alterations to unique cell types in the AD brain.

**Figure 7.**
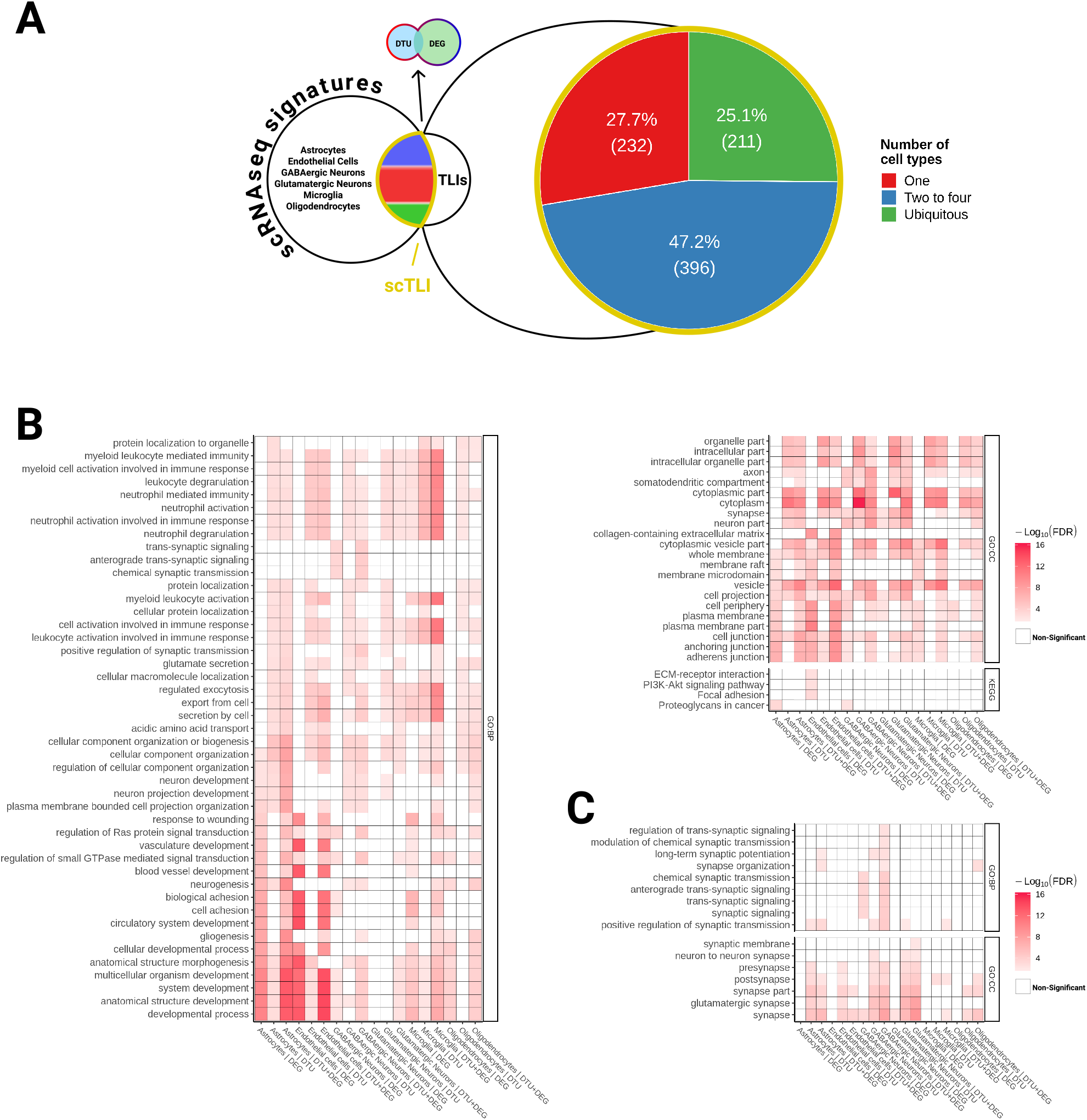
Cell-type expression pattern for genes altered in AD brains. A) Schematic representation showing our strategy to assign DEGs and gDTUs identified in the TLI to specific cell-types of the adult human brain (see also Supplementary Figure 1). Out of 839 single-cell TLI genes (scTLI), 281 were expressed in a unique cell-type, 249 in 2 to 4 cell-types and 77 in all cell-types/subtypes analyzed. B) Gene ontology terms enriched for scTLI DEGs, gDTUs or both per cell type. C) Selected GO terms associated with synaptic transmission.

### DEG/gDTU analyses identify cell-type specific alterations in AD risk/causal genes

Genomic association studies have revealed about 45 loci containing variants related to an increased or decreased probability of developing AD (Kunkle et al., 2019; Lambert et al., 2013). However, the functional variants and their target genes remain mostly elusive (Dourlen et al., 2019). To contribute with the identification of target genes, we first evaluate the expression of 176 genes located within the 45 loci associated with AD AD risk (Supplementary table 10; Dourlen et al., 2019) and 3 causal AD genes – PSEN1, PSEN2 and APP - in individual cell types of the adult human brain. We found that 116 out of the 179 AD risk/causal genes were expressed by at least one of the major cell types identified in the brain (Figure 8A; Supplementary table 11). Subsets of these genes were exclusively expressed either in microglial cells (14 out of 116), neurons (12), astrocytes (2), oligodendrocytes (6) or endothelial cells (6), suggesting cell-type specific roles for these AD risk/causal genes.

**Figure 8.**
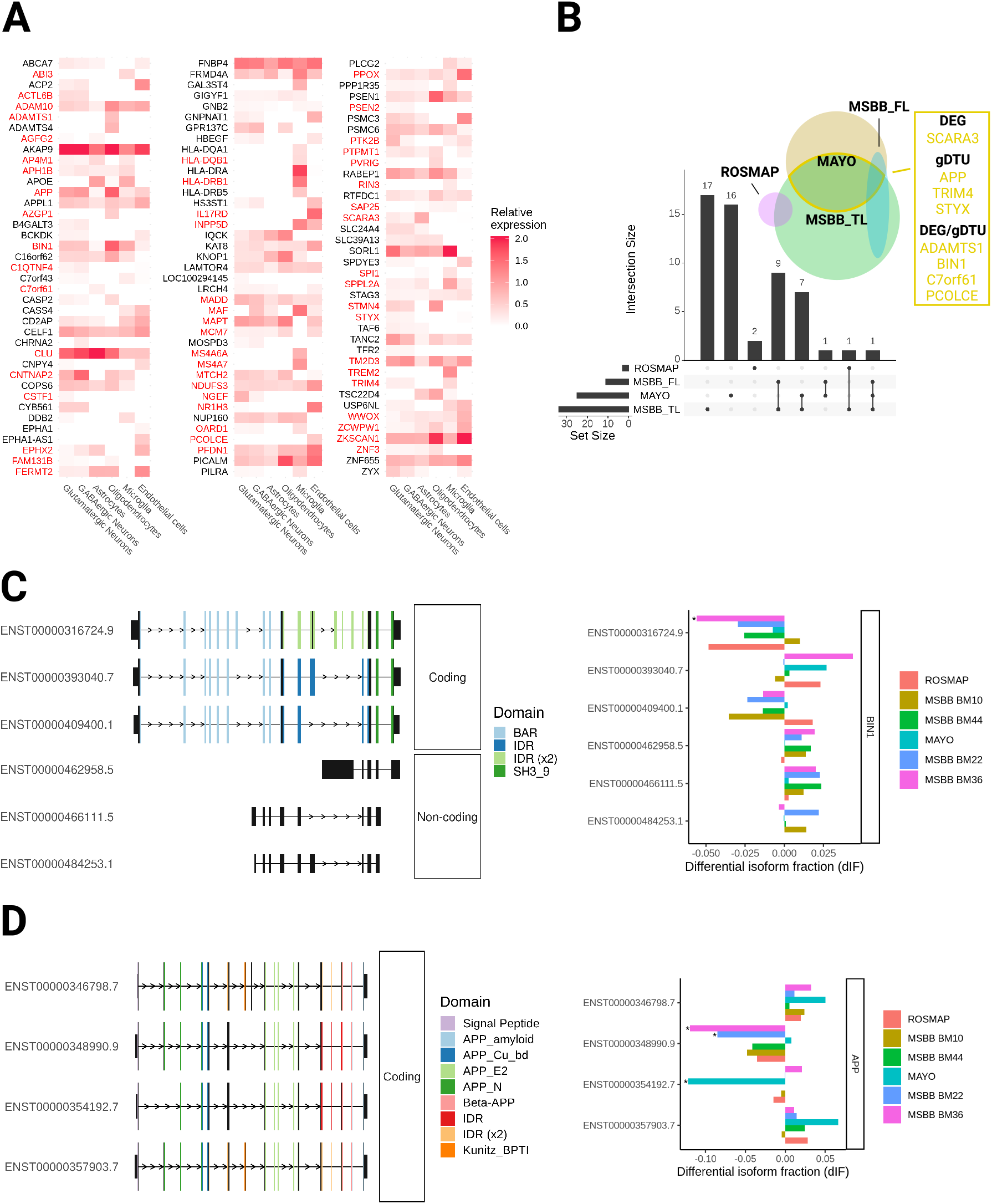
Expression of AD risk/causal genes is mostly altered in the TL of patients. A) Heatmap showing the expression of predicted AD risk/causal genes in different cell types of the adult human brain. DEGs and gDTUs in at least one dataset are highlighted in red. B) Venn diagram showing the number of AD risk/causal DEGs or gDTUs identified in the different datasets analyzed. The intersection between Mayo and MSBB TL is highlighted in yellow and genes identified are shown in the yellow box. C) Representation of the 6 most significant BIN1 isoforms altered (left) and quantification of the differential isoform fraction (dIF) in AD brains compared to controls (right). Main protein domains are indicated with different colors. D) Similar representation for APP. * dIF > 0.05 and FDR > 0.01.

Next, we set out to evaluate the differential expression or transcript usage for these genes. Out of the 116 AD risk/causal genes expressed by brain cell types (Figure 8A), we observed that 54 were also DEGs/gDTUs in at least one of the bulk RNAseq datasets analyzed. Among those genes, 2 were exclusively identified in the FL (Figure 8B). We, therefore, decided to focus on the 52 AD risk/causal genes identified in the temporal lobe for further analyses. In this region, we identified 27 and 17 DEGs/gDTUs in the MSBB_TL and Mayo datasets, respectively, including some well-characterized AD risk genes, such as ADAM10 (ADAM Metallopeptidase Domain 10), BIN1, CLU (Clusterin) and TREM2 (Triggering Receptor Expressed On Myeloid Cells 2), and the causal AD genes APP, PSEN1 and 2 (Presenilin 1 and 2) (Figure 8A-B). Eight genes were altered in both datasets (Figure 6B, yellow box; 15,38% of overlap) and were selected for further analysis of isoform switch. Using ISAR to identify the isoforms altered in the AD brains compared to controls, we observed some patterns of isoform switch that could have important functional relevance (Figure 8C and D). For instance, while BIN1 transcripts ENST00000316724.9 (NP_647593.1 - isoform 1) and ENST00000409400.1 (NP_647600.1 - isoform 9) were downregulated, transcripts ENST00000393040.7 (NP_647598.1 - isoform 6) and ENST00000462958.5, ENST0000046611.5 and ENST00000484253.1 (intron retention) were upregulated (Figure 8C). This pattern could lead to a decrease of the neuronal specific BIN1 isoform 1 expression (Zhou et al., 2014), given that retained introns are non-coding sequences. Using western blotting analysis, we confirmed this decrease of BIN1 isoform 1 protein in the frontal cortex and hippocampus of AD brain samples compared to controls (Supplementary Figure 5).

We also observed isoform switches in the AD causal gene APP with possible functional consequences in neuronal cells. While two APP isoforms were downregulated (ENST00000348990 and ENST00000354192), the isoforms ENST00000346798 and ENST00000357903 were upregulated in Mayo and MSBB datasets (Figure 8D). Noteworthy, significantly downregulated APP isoforms lack exon 7, which contains the Kunitz protease inhibitor (KPI) domain. KPI is one of the main serine protease inhibitors and increased KPI(+)APP mRNA and protein expression levels have been described in AD brains and are associated with increased amyloid beta deposition (Tanzi et al., 1989; Johnson et al., 1989; Kitaguchi al., 1988). At the exception of ENST00000354192, the other transcripts are mostly expressed in neurons (Marques-Coelho and Costa, unpublished data), indicating that these cells may have a selective increase in the expression of KPI(+)APP and, consequently, enhanced production of Aβ1-42.

## Discussion

A comprehensive knowledge of gene expression alterations associated with the onset and progression of human diseases is a key step towards the understanding of their cellular and molecular mechanisms (Lee & Young, 2013). In this work, we provide a novel framework to identify cell-type specific gene expression alterations in AD using patient-derived bulk RNAseq. Comparing RNA-sequencing data obtained from distinct brain regions of control and AD patients, we show that changes in gene expression are more significant in the temporal than frontal lobe. We also show that a large number of genes present isoform switches without changes in the global expression levels. As a consequence, these genes are overlooked in classical differential expression analysis, but can be detected through differential transcript usage analysis. Gene isoform switches are mostly evident at late stages of the pathology and correlate with altered expression of genes encoding for splicing-related proteins. Using an indirect approach to assign genes to unique cell types, we are also able to map DEGs/gDTUs to unique cell populations of the adult brain, and our results are comparable to previously published scRNAseq data (Grubman et al, 2019; Mathys et al, 2019). Finally, we show that a subset of AD causal/risk factors such as APP or BIN1 are differentially expressed in the AD brain. Altogether, our work provides a comprehensive description of regional and cell-type specific gene expression changes in the AD brain and suggests that alternative splicing could be an important mechanism for pathological progression. Despite the availability of RNAseq datasets generated from healthy subjects and AD patients (Allen et al, 2016; Wang et al, 2018; De Jager et al, 2018), a systematic evaluation of the gene expression changes in the AD brain, as well as comparisons of these changes in distinct brain region, was missing. To the best of our knowledge, only one study aimed at comparing gene expression levels in different AD brain regions (Haroutunian et al., 2009), but this work was based on microarray data which has a limited gene coverage. We here show, using bulk tissue RNAseq data, that alterations in gene expression are highly prominent in biological samples obtained from the temporal lobe, which harbors the first brain regions affected in the AD pathogenesis (Braak and Braak, 1991). Conversely, few changes are present in biological samples derived from the frontal lobe, where cells are affected only at advanced stages of the AD. These observations are in line with recent data showing that changes in protein expression levels in AD brains are much more prominent in the temporal lobe (hippocampus, entorhinal cortex and cingulate gyrus) than in the frontal lobe (motor cortex) (Xu et al., 2019). They can also help to explain the low number of DEGs identified in scRNAseq data obtained from the frontal lobe (Mathys et al., 2019) compared to a similar study in the entorhinal cortex (Grubman et al., 2019).

In order to minimize the variability in RNAseq experiments (Van den Berge et al., 2017) we here focused on DEGs (genes with FC>1.3 and FDR<0.01 in AD versus control) detected independently in at least two datasets containing samples of similar brain regions (TLI or FLI). These stringent criteria limited the number of DEGs used in subsequent analyses, but still allowed the uncovering of several genes previously associated with AD pathology, such as ABCA1, ABCA2, CALB1, C1R, C1S, GAD1/2, PVALB, REST, SLC32A1, SST, VGF and VIP (Abuznait & Kaddoumi, 2012; Beckmann et al., 2020; Benoit et al., 2013; Prévot & Sibille, 2020; Zullo et al., 2019). The reduced number of DEGs in FLI and TLI likely explains our failure to detect functional annotations associated with synaptic transmission and immune response in GSEA, as previously reported (Canchi et al., 2019). However, this study analyzed only the ROSMAP dataset and considered genes with FDR<0.05 as significant, regardless of the fold change, identifying 1,722 DEGs in AD versus control brains. Besides the questionable meaning of DEGs with very small fold changes, the use of such a large set of genes for GSEA can artificially increase the number of significantly enriched functional annotations and is not advised (Subramanian et al., 2005).

Nevertheless, our failure to detect key functional annotations associated with AD pathology while inputting TLI DEGs is puzzling and could suggest that DEG analysis fails to detect relevant alterations in gene expression in the AD brain. Indeed, classical DEG analysis using DESeq or edgeR, which rank all gene transcripts, including non-coding sequences (Costa-Silva et al., 2017), are insensitive to the dynamics of gene expression that could, for example, lead to isoform switches with important functional consequences (Vitting-Seerup and Sandelin, 2017). Therefore, important gene expression alterations could occur at the level of transcripts, without significant changes in the global expression of genes. According to this possibility, we provide convincing evidence that a high number of genes in the AD brain show isoform switches (DTU) but are not detected by DEG analysis, including several genes associated with the regulation of synapse transmission such as APP, NSG1, RELN, GABRA1 (Alberi et al., 2005; Kamenetz et al., 2003; Limon et al., 2012; Norstrom et al., 2010; Yu et al., 2016). Moreover, gDTUs identified in two independent datasets (TLI), alone or in combination with TLI DEGs, were enriched for key biological processes involved in AD pathogenesis, such as synaptic communication, immune response, inflammation, endocytosis and cell-signaling (Canter et al., 2016). Similar gene set enrichment has been described using the analysis of co-expression modules in bulk RNAseq (Milind et al., 2020; Wan et al., 2020) or DEG analysis DEGs in unique cell types in scRNAseq (Mathys et al., 2018; Grubman et al., 2018). This could suggest that the combination of DEG and DTU to analyze bulk RNAseq is comparable to scRNAseq regarding the sensitivity to detect gene expression alteration in AD brains. In agreement with this possibility, we were able to assign DEGs and gDTUs to unique cell types and confirm the similarities among cell type-specific functional annotations observed in our work compared to previous scRNAseq studies (Mathys et al., 2019; Grubman et al., 2019).

Notably, we show that several DEGs/gDTUs associated with AD pathogenesis, such as NSG1, CALB1, RELN (Alberi et al., 2005; Norstrom et al., 2010; Yu et al., 2015; Odero et al., 2010) are exclusively assigned to GABAergic neurons. These genes may be particularly relevant for AD pathogenesis, given the central role of GABAergic neurons for generation of oscillatory rhythms, network synchrony, and memory in different animal models of AD (Verret et al., 2012; Etter et al., 2019). Isoform switches in the APP gene could particularly affect GABAergic neurons, which express high levels of that gene, contributing to AD pathogenesis. According to this possibility, conditional knockout of APP APP/APLP2 only in GABAergic forebrain neurons using DlxCre mice leads to cognitive deficits in hippocampus-dependent spatial learning and memory tasks, associated with altered neuronal morphology and synaptic plasticity (Mehr et al., 2020). It is tempting to speculate that GABAergic neurons could be particularly vulnerable in AD, contributing to the increased neuronal activity and synapse downscaling observed in AD brains (Canter et al., 2016; Dörrbaum et al., 2020).

The high number of gDTUs observed in AD brains compared to controls can likely be explained by altered expression of genes encoding for proteins of the splicing machinery, affecting alternative splicing. According to this interpretation, we show that a high number of isoform switches is associated with alternative transcription start site, alternative transcription termination site, exon skipping, alternative 3’ or 5’ splice sites, multiple exon skipping and intron retention. Moreover, we show that several genes encoding for proteins of the splicing machinery have their expression altered in AD brains, especially those showing a high degree of pathology (Braak > 5). Also in agreement with the regional differences in gene expression described above, alterations in the splicing machinery are more prominent in the TL than in the FL, which could help to explain the low number of gDTUs in the latter brain region identified in our work and in previous study using a different strategy to detect isoform switch (Raj et al., 2018).

Particularly interesting, several genes encoding for proteins involved in the control of alternative splicing in neurons are differently expressed in the TL of AD brains. For instance, RBFOX1 and 2 are down regulated in the MSBB BM36 and could contribute to the altered rate of exon skipping observed in this region (Alam et al., 2014; Raj & Blencowe, 2015). Noteworthy, reduced expression of RBFOX1 has been associated with an increased inclusion of exon 7 in the APP gene, leading to an enhanced expression of APP isoforms 770 and 751 containing the KPI domain (Alam et al., 2014). A similar switch in the APP isoforms has also been associated with somatic gene recombination in AD (M. H. Lee et al., 2018), indicating that increased ratios of APP isoforms containing the KPI domain could be detrimental to neurons. Considering these findings and the well-established associations between APP-KPI expression levels, amyloid plaque deposition and AD pathology progression (Tanzi et al., 1988, Johnson et al. 1990; Kitaguchi et al., 1988), it is tempting to speculate that controlling APP isoform switches by manipulating RBFOX family proteins could be a potential therapeutic strategy to hamper disease progression.

Altered exon skipping could also help to explain the isoform switch observed for BIN1, which is a major risk factor for AD (Lambert et al., 2013; Kunkle et al., 2019). BIN1 comprises a N-BAR domain involved in membrane curvature sensing, an SH3 domain that binds to proline-rich motifs, and a clathrin-binding domain (CLAP) specific of the neuronal isoform 1 (Zhou et al., 2014). We show that the transcript encoding for this latter isoform is significantly reduced in the temporal lobe, suggesting that expression of BIN1 isoform 1 in neurons is reduced. This observation is in line with decreased BIN1 isoform 1 protein expression in the AD brain compared with controls (our own results; Glennon et al., 2013). This would be also in agreement with the observation that an overexpression of the BIN1 isoform 1 may be protective in a model of Tauopathy (Sartori et al., 2019).

Although we cannot formally rule out that a stage-dependent increase in the number of DEGs and gDTUs could be due to the loss of neuronal cells in brain regions affected by the pathology, several lines of evidence indicate that this is not the most parsimonious explanation for the data described here. Firstly, we observe that the percentage of up and down regulated genes in GABAergic and glutamatergic neurons are close to 50%, ruling out the possibility that changes in cell numbers could explain these changes. Secondly, previous scRNAseq studies in AD observed a consistent fraction of cell types isolated across control and AD individuals (Mathys et al., 2019; Grubman et al., 2019), ruling out significant changes in cellular composition of AD brains. Lastly, the large number of genes with total expression levels unchanged but presenting isoform switches in the AD brains may likely presuppose a steady cellular composition of the tissue.

Altogether, our work proposes a novel strategy to analyze bulk RNAseq data and identify meaningful gene expression alterations in the diseased brain. It also corroborates previous work implicating alternative splicing in AD susceptibility (Raj et al., 2018) and suggests that isoform switches in the gene BIN1 are involved in the reduced expression of the main neuronal BIN1 isoform 1 in AD brains.

## Methods

### Bulk RNAseq data from human control and disease banks

RNAseq datasets obtained from different brain regions were used (Mayo: Allen et al, 2016; MSBB: Wang et al, 2018; ROSMAP: De Jager et al, 2018). Datasets were downloaded from AMP-AD Knowledge Portal (https://www.synapse.org) following all terms and conditions to use the data. The brain area analyzed and the number of individuals per condition were the following: Mayo - Temporal cortex, which neuroanatomically subdivides into the inferior, middle, and superior temporal gyri (STG), and cytoarchitectonically can be subdivided into Brodmann areas (BM, instead of BA) 20/21/22/41/42 (Strotzer, 2009), N=160 subjects (82 AD and 78 controls); MSBB - BM22, which is part of the Wernicke’s area in the STG, N=159 subjects (98 AD and 61 controls); MSBB BM36, corresponding to the lateral perirhinal cortex, N=154 subjects (88 AD and 64 controls); MSBB BM10, corresponding to the anterior prefrontal cortex, N=176 subjects (105 AD and 71 controls); MSBB BM44, corresponding to the inferior frontal gyrus, N=153 subjects (90 AD and 63 controls); and ROSMAP - Dorsolateral prefrontal cortex (DLPFC), containing BM46 and part of BM9, N=423 subjects (222 AD and 201 controls). Unless stated otherwise, data obtained from different analyses were grouped in “temporal lobe” (TL) - Mayo, MSBB BM22 and MSBB BM26; or “frontal lobe” (FL) - ROSMAP, MSBB BM10 and MSBB BM44.

Metadata obtained from each study was used to classify patients into Control and Alzheimer’s disease groups (Supplementary table 12). Briefly, for the MSBB dataset, we used patients with Neuropathology Category (NP.1) labeled as “Control” and “definitive Alzheimer”. For the Mayo dataset, we used the “Diagnosis” column of the metadata, selecting only “AD” and “Control” patients. For the ROSMAP dataset, we also used the column “Diagnosis” of the metadata, selecting only “Control” (value = 1) and “Alzheimer with no other conditions” (value = 4). In all those datasets, subjects marked as “AD” showed Braak stage values higher than 4. In the MSBB dataset, CDR scores of AD patients were consistently higher than 2. In the Mayo and ROSMAP datasets, all AD patients had also a definitive diagnosis according to NINCDS criteria. Co-variates such as “Post-mortem interval (PMI)”, “RNA integrity number (RIN)”, “Age of death” and “Sex” were balanced among the different groups (Table 1; Chi-square test, p>0.05). We used RIN and PMI as covariates in our model to control for possible “batch effects” (linear regression). For detailed information of all individual samples used in this study, please refer to Supplementary table 13.

**Table 1.**
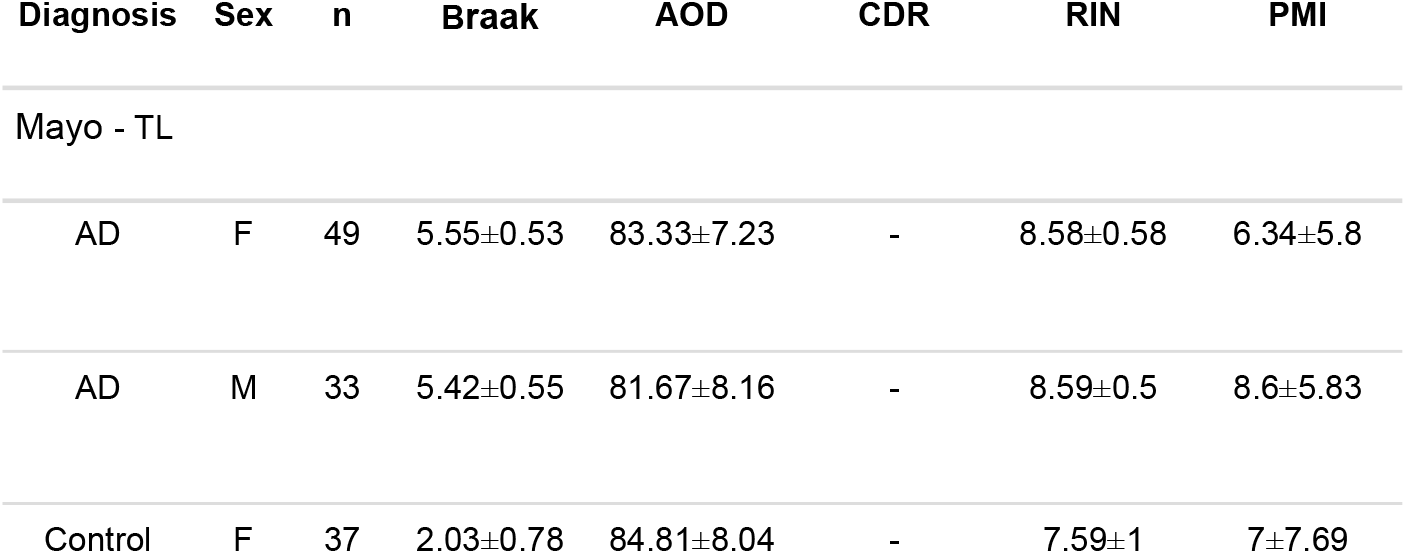
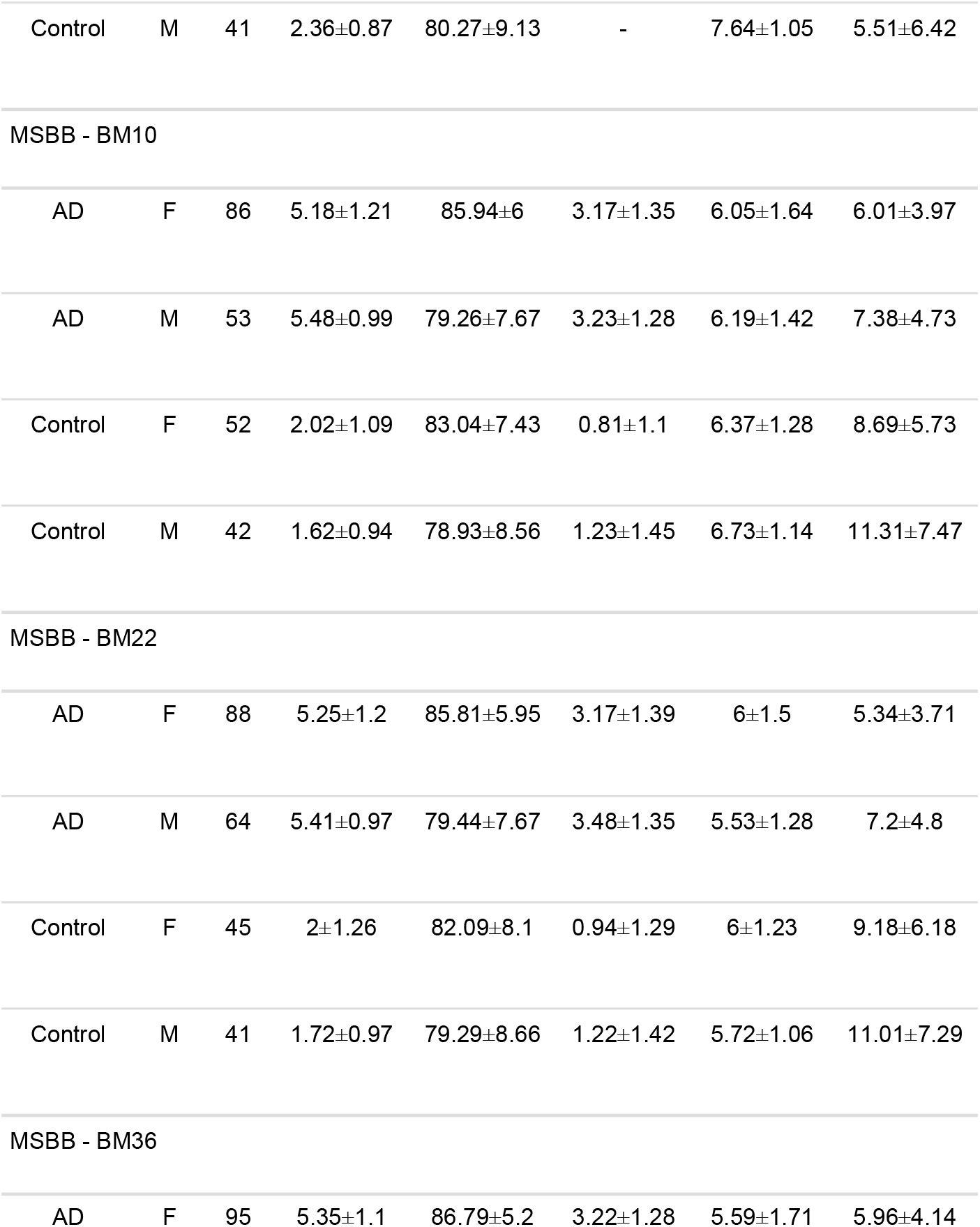
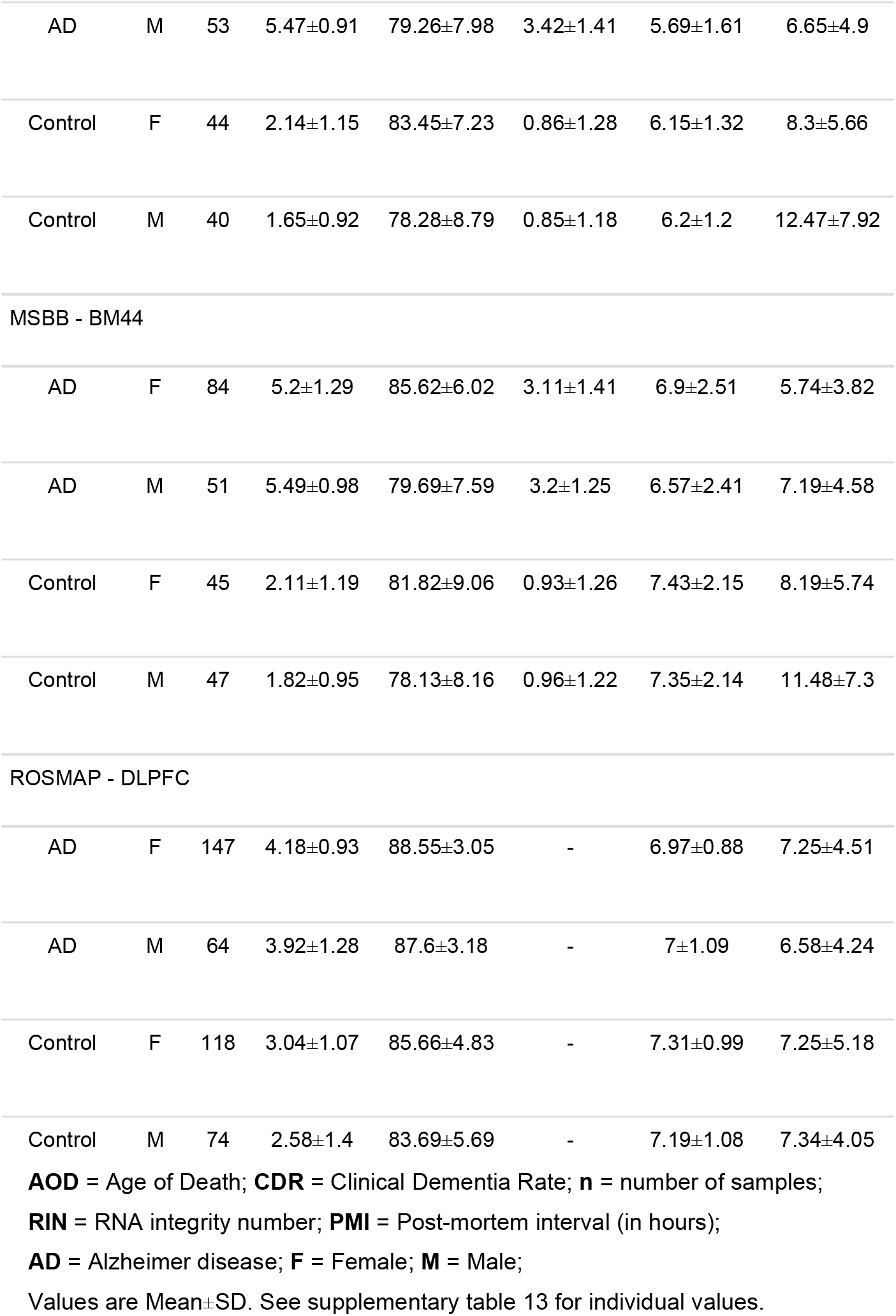
Summary of clinical, demographic and technical variables of samples analyzed from different datasets.

| Diagnosis | Sex | n | Braak | AOD | CDR | RIN | PMI |
| --- | --- | --- | --- | --- | --- | --- | --- |
| Mayo - TL |  |  |  |  |  |  |  |
| AD | F | 49 | 5.55±0.53 | 83.33±7.23 | - | 8.58±0.58 | 6.34±5.8 |
| AD | M | 33 | 5.42±0.55 | 81.67±8.16 | - | 8.59±0.5 | 8.6±5.83 |
| Control | F | 37 | 2.03±0.78 | 84.81±8.04 | - | 7.59±1 | 7±7.69 |
| Control | M | 41 | 2.36±0.87 | 80.27±9.13 | - | 7.64±1.05 | 5.51±6.42 |

|  |
| --- |
| MSBB - BM10 |
| AD | F | 86 | 5.18±1.21 | 85.94±6 | 3.17±1.35 | 6.05±1.64 | 6.01±3.97 |
| AD | M | 53 | 5.48±0.99 | 79.26±7.67 | 3.23±1.28 | 6.19±1.42 | 7.38±4.73 |
| Control | F | 52 | 2.02±1.09 | 83.04±7.43 | 0.81±1.1 | 6.37±1.28 | 8.69±5.73 |
| Control | M | 42 | 1.62±0.94 | 78.93±8.56 | 1.23±1.45 | 6.73±1.14 | 11.31±7.47 |

|  |
| --- |
| MSBB - BM22 |
| AD | F | 88 | 5.25±1.2 | 85.81±5.95 | 3.17±1.39 | 6±1.5 | 5.34±3.71 |
| AD | M | 64 | 5.41±0.97 | 79.44±7.67 | 3.48±1.35 | 5.53±1.28 | 7.2±4.8 |
| Control | F | 45 | 2±1.26 | 82.09±8.1 | 0.94±1.29 | 6±1.23 | 9.18±6.18 |
| Control | M | 41 | 1.72±0.97 | 79.29±8.66 | 1.22±1.42 | 5.72±1.06 | 11.01±7.29 |

|  |
| --- |
| MSBB - BM36 |
| AD | F | 95 | 5.35±1.1 | 86.79±5.2 | 3.22±1.28 | 5.59±1.71 | 5.96±4.14 |
| AD | M | 53 | 5.47±0.91 | 79.26±7.98 | 3.42±1.41 | 5.69±1.61 | 6.65±4.9 |
| Control | F | 44 | 2.14±1.15 | 83.45±7.23 | 0.86±1.28 | 6.15±1.32 | 8.3±5.66 |
| Control | M | 40 | 1.65±0.92 | 78.28±8.79 | 0.85±1.18 | 6.2±1.2 | 12.47±7.92 |
| MSBB - BM44 |  |  |  |  |  |  |  |
| AD | F | 84 | 5.2±1.29 | 85.62±6.02 | 3.11±1.41 | 6.9±2.51 | 5.74±3.82 |
| AD | M | 51 | 5.49±0.98 | 79.69±7.59 | 3.2±1.25 | 6.57±2.41 | 7.19±4.58 |
| Control | F | 45 | 2.11±1.19 | 81.82±9.06 | 0.93±1.26 | 7.43±2.15 | 8.19±5.74 |
| Control | M | 47 | 1.82±0.95 | 78.13±8.16 | 0.96±1.22 | 7.35±2.14 | 11.48±7.3 |
| ROSMAP - DLPFC |  |  |  |  |  |  |  |
| AD | F | 147 | 4.18±0.93 | 88.55±3.05 | - | 6.97±0.88 | 7.25±4.51 |
| AD | M | 64 | 3.92±1.28 | 87.6±3.18 | - | 7±1.09 | 6.58±4.24 |
| Control | F | 118 | 3.04±1.07 | 85.66±4.83 | - | 7.31±0.99 | 7.25±5.18 |
| Control | M | 74 | 2.58±1.4 | 83.69±5.69 | - | 7.19±1.08 | 7.34±4.05 |

**AOD** = Age of Death; **CDR** = Clinical Dementia Rate; **n** = number of samples;
**RIN** = RNA integrity number; **PMI** = Post-mortem interval (in hours);
**AD** = Alzheimer disease; **F** = Female; **M** = Male;
Values are Mean ± SD. See supplementary table 13 for individual values.

For single-cell RNA sequencing (scRNAseq) analyses, we used processed data obtained from the middle temporal gyrus (MTG), available at the Allen Brain Atlas consortium (https://celltypes.brain-map.org/rnaseq).

### Realignment of human reads into single pseudo aligner pipeline

Using human GRCh38 cDNA release 94 (ftp://ftp.ensembl.org/pub/release-94) as reference, we built an index to align all our fastq files. Next, we used pseudoaligner Kallisto (Bray et al., 2016; version 0.43.1) with our pre-built index to align fastq files.

### Differential gene expression analyses

Differentially expressed genes (DEGs) were identified using differential gene expression at transcript-level using DESeq2 R library (Love et al., 2014, 2019). To facilitate kallisto output import, transcript-level estimated counts, length and abundance was extracted using *tximport* function (Soneson et al., 2016). As described by Michael Love group, transcript-level differential gene expression enhances analysis resolution (Love et al., 2019). Using *DESeqDataSetFromTximport*, a DESeq2 object was created and filtered using rows with sum of all counts bigger than 10. Next, *DESeq* function was used with default values. Using the *results* function, we selected all genes with False Discovery Rate (FDR) < 0.01 and Fold Change (FC) > 1.3. We also used RIN and PMI as covariates (linear regression).

Differential Transcript Usage (DTU) analysis was performed using the R library IsoformSwitchAnalyzeR (Vitting-Seerup & Sandelin, 2019). Following pipeline instructions, kallisto abundance tables were imported using *importIsoformExpression* and *importRdata* functions to create a *switchAnalyzeRlist* object. Same cDNA release used in kallisto alignment and correspondent annotation (ftp://ftp.ensembl.org/pub/release-94/gtf/homo_sapiens/Homo_sapiens.GRCh38.94.chr_patch_hapl_scaff.gtf.gz) were applied as input. We filtered data using a gene expression cut off = 10, isoform expression cut off = 3, differential isoform fraction (dIF) cut off = 0.05 and removed single isoform genes. Although DEXSeq is recommended to test differential isoform usage, it does not work efficiently for large datasets (more than 100 samples; (Anders et al., 2012)). For that reason, we chose *isoformSwitchAnalysisPart1* function using DRIMSeq (Soneson et al., 2016) to test differential transcript usage. Using part1 fasta files, all external analysis was performed and used as input to *isoformSwitchAnalysisPart2* function. We used CPC2, Pfam, SignalIP and Netsurfp2 as indicated in the pipeline. Next, we performed a confirmation stage using stageR (Van den Berge et al., 2017) to generate isoforms overall false discovery rate (OFDR). We selected all isoforms with OFDR < 0.01 and dIF > 0.05. RIN and PMI metadata were used as covariates (linear regression).

Statistical significance of the intersections among different datasets was calculated using the hypergeometric test (phyper).

### Splicing events and event consequences

We used extractSplicingSummary and extractConsequenceSummary functions to quantify gain/loss of predicted splicing events (such as exon skipping and intron-retention); and gain/loss (also sensitive/insensitive, shorter/longer and switch) of predicted functional consequences (such as coding potential and domain identified), respectively.

### Single-cell RNAseq

Using R library *seurat*, we created a seurat object (*CreateSeuratObject*), normalized data (*NormalizeData*), found variable genes (*FindVariableFeatures*) and rescaled data using a linear model (*ScaleData, use*.*umi = F*). After that, we generated 50 PC’s (*RunPCA*) but only used 35 of them based on the PC’s visualization distribution (*ElbowPlot*). Since Allen data was already annotated, we only used tSNE (*RunTSNE*) to facilitate visualization. A group classified as “None” by Allen metadata were removed from our analysis. This strategy generated 7 main different cell types: Astrocytes, Endothelial cells, Glutamatergic Neurons, GABAergic Neurons, Microglia, Oligodendrocytes and oligodendrocyte precursor cells (OPCs). To assign genes to specific cell-types, we used the AverageExpression function. Using pct.exp bigger than 0.1, we created a list of genes that were expressed by each cell type.

### Gene-set enrichment analysis (GSEA)

For gene ontology analysis, R library *gprofiler2* was used. Using *gost* function, correction_method=“fdr’’ and significant=TRUE. To minimize the enrichment of gene ontologies based on small set of genes, we used three conditions for significance assessment: false discovery rate (FDR)<0.01; intersection size (intersection between gene set vs. number of genes in a term)>3; and precision (intersection size divided by gene set)>0.03. We used Gene Ontology (GO or by branch GO:MF, GO:BP, GO:CC), Kyoto Encyclopedia of Genes and Genomes (KEGG), Reactome (REAC), WikiPathways (WP), TRANSFAC (TF), miRTarBase (MIRNA), Human Protein Atlas (HPA), CORUM (CORUM), Human phenotype ontology (HP) as sources. For improved visualization, we plotted results only for GO:BP, GO:CC and KEGG and show only FDR for terms reaching all criteria of significance.

### Selection of splicing-associated genes

To select splicing related genes, we searched for terms containing the words “splicing” or “spliceosome” in gProfiler bank (https://biit.cs.ut.ee/gprofiler/gost). Taking only GO and WP datasets, 25 terms and 441 genes related to those terms were selected (Supplementary table 6).

### Selection of AD risk/causal genes

The complete list of AD risk/causal genes used in this study is described in the Supplementary Table 10. Briefly, AD risk loci were selected from previous work using Genome-wide Association Studies and Whole Exome Sequencing (Lambert et al., 2013; Kunkle et al., 2019). AD risk genes within these loci were determined based on regional association plots, assuming that the functional risk variants are located in the vicinity of the SNP producing the top signal and taking into account the linkage disequilibrium patterns and the recombination peaks within the loci of interest (Dourlen et al., 2019). Early-onset AD causal genes used in this study are APP, PSEN1 and PSEN2.

### Western blotting

Frozen brain samples obtained from the frontal cortex (FCx) and hippocampus (Hip) of 3 non-pathology (Age: 80.33 ± 3.78 years; Braak: 2.66 ± 1.15; PMI: 37.33 ± 22.50 hours) and 6 AD patients (Age: 79.57 ± 6.70 years; Braak: 6; PMI: 26.57 ± 13.40 hours) were lysed with RIPA buffer and sonicated at 100% during 10 seconds before use for the Western blotting analyses. The controls for BIN1 isoforms 1 (Iso1) and 9 (Iso9) were obtained using HEK cells transiently transfected with 1µg/ml DNA solution containing plasmids encoding for BIN1 isoforms mixed with the transfection reagent FuGENE HD (Promega) at the ratio 1:3. Cells were lysed using RIPA buffer 48h after transfection and frozen for further analyses.

Protein quantification was performed using the BCA protein assay (Thermo Scientific). 10–20 μg of total protein from extracts were separated in SDS–polyacrylamide gels 4-12% (NuPAGE Bis-Tris, Thermo Scientific) and transferred to nitrocellulose membranes (Bio-Rad). Next, membranes were incubated in milk (5% in Tris-buffered saline with 0.1% Tween-20, TTBS; 1h at RT) to block non-specific binding sites during 30min at RT, followed by several washes with TTBS. Immunoblotting was carried out with primary antibodies anti-BIN1 (Abcam, ab182562), Anti-β-ACTIN (Sigma-Aldrich, A1978) and anti-GAPDH (Millipore, AB2302) for 1h at RT on 20 RPM. The membranes were washed 3 times in TTBS, followed by incubation with HRP-conjugated secondary antibodies (Jackson, anti-Mouse 115-035-003 and anti-Rabbit 111-035-003; Thermo Scientific, anti-chicken A16054) overnight at 4°C on 20 RPM agitation. Immunoreactivity was revealed using the ECL chemiluminescence system (SuperSignal, Thermo Scientific) and imaged using the Amersham Imager 600 (GE Life Sciences). Optical densities of bands were quantified using “Gel Analyzer” plugin in Fiji (Schindelin et al., 2012).

## Data Availability

All data referred to in the manuscript is publicly available

https://doi.org/10.7303/syn3388564

https://doi.org/10.7303/syn3157743

https://doi.org/10.7303/syn5550404

## Data availability

MSBB RNAseq (De Jager et al, 2018) - https://doi.org/10.7303/syn3388564

ROSMAP RNAseq (Wang et al, 2018) - https://doi.org/10.7303/syn3157743

MAYO RNAseq (Allen et al, 2018) - https://doi.org/10.7303/syn5550404

## Code availability

Codes used for analyses are available, differentially expressed genes and differential transcript usage in individual datasets can be assessed using https://diegomscoelho.github.io/AD-IsoformSwitch/

## Acknowledgments

We thank Charles Duyckaerts (Neuroceb, GIE Neuro-CEB BB-0033-00011) for providing the brain samples. We would also like to thank Dr. Tarciso Velho, Pierre Dourlen and Fabien Delahaye for reading and suggestions on the manuscript. The results published here are in whole or in part based on data obtained from the AMP-AD Knowledge Portal (https://adknowledgeportal.synapse.org/). Study data were provided by the following sources: The Mayo Clinic Alzheimer’s Disease Genetic Studies, led by Dr. Nilufer Taner and Dr. Steven G. Younkin, Mayo Clinic, Jacksonville, FL using samples from the Mayo Clinic Study of Aging, the Mayo Clinic Alzheimer’s Disease Research Center, and the Mayo Clinic Brain Bank. Data collection was supported through funding by NIA grants P50 AG016574, R01 AG032990, U01 AG046139, R01 AG018023, U01 AG006576, U01 AG006786, R01 AG025711, R01 AG017216, R01 AG003949, NINDS grant R01 NS080820, CurePSP Foundation, and support from Mayo Foundation. Study data includes samples collected through the Sun Health Research Institute Brain and Body Donation Program of Sun City, Arizona. The Brain and Body Donation Program is supported by the National Institute of Neurological Disorders and Stroke (U24 NS072026 National Brain and Tissue Resource for Parkinson’s Disease and Related Disorders), the National Institute on Aging (P30 AG19610 Arizona Alzheimer’s Disease Core Center), the Arizona Department of Health Services (contract 211002, Arizona Alzheimer’s Research Center), the Arizona Biomedical Research Commission (contracts 4001, 0011, 05-901 and 1001 to the Arizona Parkinson’s Disease Consortium) and the Michael J. Fox Foundation for Parkinson’s Research. Study data were also provided by the Rush Alzheimer’s Disease Center, Rush University Medical Center, Chicago. Data collection was supported through funding by NIA grants P30AG10161 (ROS), R01AG15819 (ROSMAP; genomics and RNAseq), R01AG17917 (MAP), the Illinois Department of Public Health (ROSMAP), and the Translational Genomics Research Institute (genomic). For ROSMAP, additional phenotypic data can be requested at www.radc.rush.edu. And part based on data generated from postmortem brain tissue collected through the Mount Sinai VA Medical Center Brain Bank and were provided by Dr. Eric Schadt from Mount Sinai School of Medicine.

This work was co-funded by the European Union under the European Regional Development Fund (ERDF) and by the Hauts de France Regional Council (contrat n°18006176), the MEL (contract_2016_ESR_05), and the French State (contract n°2018-3-CTRL_IPL_Phase2) to MRC. This work was also funded by the Lille Métropole Communauté Urbaine and the French government’s LABEX DISTALZ program (Development of innovative strategies for a transdisciplinary approach to Alzheimer’s disease) to JCL. DMC and LICC are supported by Coordenaç ão de Aperfeiçoamento de Pessoal de Nível Superior (CAPES) scholarships.

## Author Contributions

M.R.C. conceived of the presented idea and supervised the experiments. D.M-C. performed all bioinformatic analyses and prepared the figures and tables of the manuscript. L.I.C.C. contributed with bioinformatic analyses. A.R.M.F. performed western blot analysis in human samples. D.M-C., J-C. L. and M.R.C. wrote the manuscript. All authors revised and approved the final version of the manuscript.

## References

Abuznait, A. H., & Kaddoumi, A. (2012). Role of ABC transporters in the pathogenesis of Alzheimers disease. ACS Chemical Neuroscience, 3(11), 820–831. https://doi.org/10.1021/cn300077c

Alam, S., Suzuki, H., & Tsukahara, T. (2014). Alternative splicing regulation of APP exon 7 by RBFox proteins. Neurochemistry International, 78, 7–17. https://doi.org/10.1016/j.neuint.2014.08.001

Alberi, S., Boda, B., Steiner, P., Nikonenko, I., Hirling, H., & Muller, D. (2005). The endosomal protein NEEP21 regulates AMPA receptor-mediated synaptic transmission and plasticity in the hippocampus. Molecular and Cellular Neuroscience, 29(2), 313–319. https://doi.org/10.1016/j.mcn.2005.03.011

Allen, M., Carrasquillo, M. M., Funk, C., Heavner, B. D., Zou, F., Younkin, C. S., Burgess, J. D., Chai, H. S., Crook, J., Eddy, J. A., Li, H., Logsdon, B., Peters, M. A., Dang, K. K., Wang, X., Serie, D., Wang, C., Nguyen, T., Lincoln, S., … Ertekin-Taner, N. (2016). Human whole genome genotype and transcriptome data for Alzheimer’s and other neurodegenerative diseases. Scientific Data, 3. https://doi.org/10.1038/sdata.2016.89

Anders, S., Reyes, A., & Huber, W. (2012). Detecting differential usage of exons from RNA-seq data. Genome Research, 22(10), 2008–2017. https://doi.org/10.1101/gr.133744.111

Arzalluz-Luqueángeles, & Conesa, A. (2018). Single-cell RNAseq for the study of isoforms-how is that possible? Genome Biology, 19(1), 1–19. https://doi.org/10.1186/s13059-018-1496-z

Braak, H., & Braak, E. (1991). Neuropathological stageing of Alzheimer-related changes. Acta Neuropathol (Vol. 82): 239–259

Beckmann, N. D., Lin, W., Wang, M., Cohain, A. T., Charney, A. W., Wang, P., Ma, W., Wang, Y., Jiang, C., Audrain, M., Comella, P. H., Fakira, A. K., Hariharan, S. P., Belbin, G. M., Girdhar, K., Levey, A. I., Seyfried, N. T., & Dammer, E. B. (2020). regulator of Alzheimer’ s disease. Nature Communications. https://doi.org/10.1038/s41467-020-17405-z

Benoit, M. E., Hernandez, M. X., Dinh, M. L., Benavente, F., Vasquez, O., & Tenner, A. J. (2013). C1q-induced LRP1B and GPR6 proteins expressed early in Alzheimer disease mouse models, are essential for the C1q-mediated protection against amyloid-β neurotoxicity. Journal of Biological Chemistry, 288(1), 654–665. https://doi.org/10.1074/jbc.M112.400168

Bray, N. L., Pimentel, H., Melsted, P., & Pachter, L. (2016). Near-optimal probabilistic RNA-seq quantification. Nature Biotechnology, 34(5), 525–527. https://doi.org/10.1038/nbt.3519

Canchi, S., Raao, B., Masliah, D., Rosenthal, S. B., Sasik, R., Fisch, K. M., De Jager, P. L., Bennett, D. A., & Rissman, R. A. (2019). Integrating Gene and Protein Expression Reveals Perturbed Functional Networks in Alzheimer’s Disease. Cell Reports, 28(4), 1103-1116.e4. https://doi.org/10.1016/j.celrep.2019.06.073

Canter, R. G., Penney, J., & Tsai, L. H. (2016). The road to restoring neural circuits for the treatment of Alzheimer’s disease. In Nature (Vol. 539, Issue 7628,pp. 187–196). Nature Publishing Group. https://doi.org/10.1038/nature20412

Choi, J. H. K., Kaur, G., Mazzella, M. J., Morales-Corraliza, J., Levy, E., & Mathews, P. M. (2013). Early endosomal abnormalities and cholinergic neuron degeneration in Amyloid-β protein precursor transgenic mice. Journal of Alzheimer’s Disease, 34(3), 691–700. https://doi.org/10.3233/JAD-122143

Costa-Silva, J., Domingues, D., & Lopes, F. M. (2017). RNA-Seq differential expression analysis: An extended review and a software tool. PLoS ONE, 12(12), 1–18. https://doi.org/10.1371/journal.pone.0190152

De Jager, P. L., Ma, Y., McCabe, C., Xu, J., Vardarajan, B. N., Felsky, D., Klein, H. U., White, C. C., Peters, M. A., Lodgson, B., Nejad, P., Tang, A., Mangravite, L. M., Yu, L., Gaiteri, C., Mostafavi, S., Schneider, J. A., & Bennett, D. A. (2018). Data descriptor: A multi-omic atlas of the human frontal cortex for aging and Alzheimer’s disease research. Scientific Data, 5. https://doi.org/10.1038/sdata.2018.142

Devi, L., & Ohno, M. (2015). TrkB reduction exacerbates Alzheimer’s disease-like signaling aberrations and memory deficits without affecting β-amyloidosis in 5XFAD mice. Translational Psychiatry, 5(5), e562–9. https://doi.org/10.1038/tp.2015.55

Dörrbaum, A.R., Alvarez-Castelao, B., Nassim-Assir, B., Langer, J. D., & Schuman, E. M. (2020). Proteome dynamics during homeostatic scaling in cultured neurons. Elife e2020;9:e52939. DOI: https://doi.org/10.7554/eLife.52939

Dourlen, P., Kilinc, D., Malmanche, N., Chapuis, J., & Lambert, J. C. (2019). The new genetic landscape of Alzheimer’s disease: from amyloid cascade to genetically driven synaptic failure hypothesis? In Acta Neuropathologica (Vol. 138, Issue 2, pp. 221–236). Springer Verlag. https://doi.org/10.1007/s00401-019-02004-0

Glennon, E. B. C., Whitehouse, I. J., Miners, J. S., Kehoe, P. G., Love, S., Kellett, K. A. B., & Hooper, N. M. (2013). BIN1 Is Decreased in Sporadic but Not Familial Alzheimer’s Disease or in Aging. PLoS ONE, 8(10), 1–11. https://doi.org/10.1371/journal.pone.0078806

Grubman, A., Chew, G., Ouyang, J. F., Sun, G., Choo, X. Y., McLean, C., Simmons, R. K., Buckberry, S., Vargas-Landin, D. B., Poppe, D., Pflueger, J., Lister, R., Rackham, O. J. L., Petretto, E., & Polo, J. M. (2019). A single-cell atlas of entorhinal cortex from individuals with Alzheimer’s disease reveals cell-type-specific gene expression regulation. Nature Neuroscience, 22(12), 2087–2097. https://doi.org/10.1038/s41593-019-0539-4

Haroutunian, V., Katsel, P., & Schmeidler, J. (2009). Transcriptional vulnerability of brain regions in Alzheimer’s disease and dementia. Neurobiology of Aging, 30(4), 561–573. https://doi.org/10.1016/j.neurobiolaging.2007.07.021

Johnson, S. A., Rogers, J., & Finch, C. E. (1989). APP-695 transcript prevalence is selectively reduced during Alzheimer’s disease in cortex and hippocampus but not in cerebellum. In Neurobiology of Aging (Vol. 10, Issue 6). ∼ Pergamon Press plc. https://doi.org/10.1016/0197-4580(89)90017-1

Kamenetz, F., Tomita, T., Hsieh, H., Seabrook, G., Borchelt, D., Iwatsubo, T., Sisodia, S., & Malinow, R. (2003). APP Processing and Synaptic Function. Neuron, 37(6), 925–937. https://doi.org/10.1016/S0896-6273(03)00124-7

Kunkle, B. W., Grenier-Boley, B., Sims, R., Bis, J. C., Damotte, V., Naj, A. C., Boland, A., Vronskaya, M., van der Lee, S. J., Amlie-Wolf, A., Bellenguez, C., Frizatti, A., Chouraki, V., Martin, E. R., Sleegers, K., Badarinarayan, N., Jakobsdottir, J., Hamilton-Nelson, K. L., Moreno-Grau, S., … Pericak-Vance, M. A. (2019). Genetic meta-analysis of diagnosed Alzheimer’s disease identifies new risk loci and implicates Aβ, tau, immunity and lipid processing. Nature Genetics, 51(3), 414–430. https://doi.org/10.1038/s41588-019-0358-2

Lambert, J. C., Ibrahim-Verbaas, C. A., Harold, D., Naj, A. C., Sims, R., Bellenguez, C., Jun, G., DeStefano, A. L., Bis, J. C., Beecham, G. W., Grenier-Boley, B., Russo, G., Thornton-Wells, T. A., Jones, N., Smith, A. V., Chouraki, V., Thomas, C., Ikram, M. A., Zelenika, D., … Seshadri, S. (2013). Meta-analysis of 74,046 individuals identifies 11 new susceptibility loci for Alzheimer’s disease. Nature Genetics, 45(12), 1452–1458. https://doi.org/10.1038/ng.2802

Lee, M. H., Siddoway, B., Kaeser, G. E., Segota, I., Rivera, R., Romanow, W. J., Liu, C. S., Park, C., Kennedy, G., Long, T., & Chun, J. (2018). Somatic APP gene recombination in Alzheimer’s disease and normal neurons. Nature, 563(7733), 639–645. https://doi.org/10.1038/s41586-018-0718-6

Lee, T. I., & Young, R. A. (2013). Transcriptional regulation and its misregulation in disease. In Cell. https://doi.org/10.1016/j.cell.2013.02.014

Limon, A., Reyes-Ruiz, J. M., & Miledi, R. (2012). Loss of functional GABA A receptors in the Alzheimer diseased brain. Proceedings of the National Academy of Sciences of the United States of America, 109(25), 10071–10076. https://doi.org/10.1073/pnas.1204606109

Love, M. I., Huber, W., & Anders, S. (2014). Moderated estimation of fold change and dispersion for RNA-seq data with DESeq2. Genome Biology, 15(12), 1–21. https://doi.org/10.1186/s13059-014-0550-8

Love, M. I., Soneson, C., Patro, R., Vitting-seerup, K., & Oshlack, A. (2019). Swimming downstream?: statistical analysis of differential transcript usage following Salmon quantification [version 1?; peer review?: 3 approved with reservations ] Referee Status?: This article is included in the RPackage gateway. (IssueMay).

Masnada, S., Hedrich, U. B. S., Gardella, E., Schubert, J., Kaiwar, C., Klee, E. W., Lanpher, B. C., Gavrilova, R. H., Synofzik, M., Bast, T., Gorman, K., King, M. D., Allen, N. M., Conroy, J., Ben Zeev, B., Tzadok, M., Korff, C., Dubois, F., Ramsey, K., … Rubboli, G. (2017). Clinical spectrum and genotype-phenotype associations of KCNA2-related encephalopathies. Brain, 140(9), 2337–2354. https://doi.org/10.1093/brain/awx184

Masters, C. L., Bateman, R., Blennow, K., Rowe, C. C., Sperling, R. A., & Cummings, J. L. (2015). Alzheimer’s disease. In Nature Reviews Disease Primers (Vol. 1). Nature Publishing Group. https://doi.org/10.1038/nrdp.2015.56

Mathys, H., Davila-Velderrain, J., Peng, Z., Gao, F., Mohammadi, S., Young, J. Z., Menon, M., He, L., Abdurrob, F., Jiang, X., Martorell, A. J., Ransohoff, R. M., Hafler, B. P., Bennett, D. A., Kellis, M., & Tsai, L. H. (2019). Single-cell transcriptomic analysis of Alzheimer’s disease. Nature, 570(7761), 332–337. https://doi.org/10.1038/s41586-019-1195-2

Mehr, A., Hick, M., Ludewig, S., Müller, M., Herrmann, U., von Engelhardt, J., Wolfer, D. P., Korte, M., & Müller, U. C. (2020). Lack of APP and APLP2 in GABAergic Forebrain Neurons Impairs Synaptic Plasticity and Cognition. Cerebral Cortex (New York, N.Y.?: 1991), 30(7), 4044–4063. https://doi.org/10.1093/cercor/bhaa025

Milind, N., Preuss, C., Haber, A., Ananda, G., Mukherjee, S., John, C., Shapley, S., Logsdon, B. A., Crane, P. K., & Carter, G. W. (2020). Transcriptomic stratification of late-onset Alzheimer’s cases reveals novel genetic modifiers of disease pathology. PLoS Genetics, 16(6), e1008775. https://doi.org/10.1371/journal.pgen.1008775

Mostafavi, S., Gaiteri, C., Sullivan, S. E., White, C. C., Tasaki, S., Xu, J., Taga, M., Klein, H. U., Patrick, E., Komashko, V., McCabe, C., Smith, R., Bradshaw, E. M., Root, D. E., Regev, A., Yu, L., Chibnik, L. B., Schneider, J. A., Young-Pearse, T. L.,…De Jager, P. L. (2018). A molecular network of the aging human brain provides insights into the pathology and cognitive decline of Alzheimer’s disease. Nature Neuroscience, 21(6), 811–819. https://doi.org/10.1038/s41593-018-0154-9

Norstrom, E. M., Zhang, C., Tanzi, R., & Sisodia, S. S. (2010). Identification of NEEP21 as a β-amyloid precursor protein-interacting protein in vivo that modulates amyloidogenic processing in vitro. Journal of Neuroscience, 30(46), 15677–15685. https://doi.org/10.1523/JNEUROSCI.4464-10.2010

Odero, G. L., Oikawa, K., Glazner, K. A. C., Schapansky, J., Grossman, D., Thiessen, J. D., Motnenko, A., Ge, N., Martin, M., Glazner, G. W., & Albensi, B. C. (2010). Evidence for the involvement of calbindin D28k in the presenilin 1 model of Alzheimer’s disease. Neuroscience, 169(1), 532–543. https://doi.org/10.1016/j.neuroscience.2010.04.004

Prévot, T., & Sibille, E. (2020). Altered GABA-mediated information processing and cognitive dysfunctions in depression and other brain disorders. Molecular Psychiatry. https://doi.org/10.1038/s41380-020-0727-3

Raj, B., & Blencowe, B. J. (2015). Alternative Splicing in the Mammalian Nervous System: Recent Insights into Mechanisms and Functional Roles. In Neuron (Vol. 87, Issue 1, pp. 14–27). Cell Press. https://doi.org/10.1016/j.neuron.2015.05.004

Raj, T., Li, Y. I., Wong, G., Humphrey, J., Wang, M., Ramdhani, S., Wang, Y. C., Ng, B., Gupta, I., Haroutunian, V., Schadt, E. E., Young-Pearse, T., Mostafavi, S., Zhang, B., Sklar, P., Bennett, D. A., & De Jager, P. L. (2018). Integrative transcriptome analyses of the aging brain implicate altered splicing in Alzheimer’s disease susceptibility. Nature Genetics, 50(11), 1584–1592. https://doi.org/10.1038/s41588-018-0238-1

Sartori, M., Mendes, T., Desai, S., Lasorsa, A., Herledan, A., Malmanche, N., Mäkinen, P., Marttinen, M., Malki, I., Chapuis, J., Flaig, A., Vreulx, A. C., Ciancia, M., Amouyel, P., Leroux, F., Déprez, B., Cantrelle, F. X., Maréchal, D., Pradier, L., … Lambert, J. C. (2019). BIN1 recovers tauopathy-induced long-term memory deficits in mice and interacts with Tau through Thr348 phosphorylation. Acta Neuropathologica, 138(4), 631–652. https://doi.org/10.1007/s00401-019-02017-9

Schindelin, J.; Arganda-Carreras, I. & Frise, E. et al. (2012), “Fiji: an open-source platform for biological-image analysis”, Nature methods 9(7): 676–682, PMID 22743772, https://doi:10.1038/nmeth.2019.

Soneson, C., Love, M. I., & Robinson, M. D. (2016). Differential analyses for RNA-seq: Transcript-level estimates improve gene-level inferences. F1000Research, 4, 1–18. https://doi.org/10.12688/F1000RESEARCH.7563.2

Strotzer, M. (2009). One century of brain mapping using Brodmann areas. Clinical Neuroradiology, 19(3), 179–186. https://doi.org/10.1007/s00062-009-9002-3

Subramanian, A., Tamayo, P., Mootha, V. K., Mukherjee, S., Ebert, B. L., Gillette, M. A., Paulovich, A., Pomeroy, S. L., Golub, T. R., Lander, E. S., & Mesirov, J. P. (2005). Gene set enrichment analysis: A knowledge-based approach for interpreting genome-wide expression profiles. Proceedings of the National Academy of Sciences of the United States of America, 102(43), 15545–15550. https://doi.org/10.1073/pnas.0506580102

Tanzi, R. E., McClatchey, A. I., Lamperti, E. D., Villa-Komaroff, L., Gusella, J. F., & Neve, R. L. (1988). Protease inhibitor domain encoded by an amyloid protein precursor mRNA associated with Alzheimer’s disease. In Nature (Vol. 331, Issue 6156). https://doi.org/10.1038/331528a0

Van den Berge, K., Soneson, C., Robinson, M. D., & Clement, L. (2017). stageR: A general stage-wise method for controlling the gene-level false discovery rate in differential expression and differential transcript usage. Genome Biology, 18(1), 1–14. https://doi.org/10.1186/s13059-017-1277-0

Verret, L., Mann, E. O., Hang, G. B., Barth, A. M. I., Cobos, I., Ho, K., Devidze, N., Masliah, E., Kreitzer, A. C., Mody, I., Mucke, L., & Palop, J. J. (2012). Inhibitory interneuron deficit links altered network activity and cognitive dysfunction in alzheimer model. Cell, 149(3), 708–721. https://doi.org/10.1016/j.cell.2012.02.046

Vitting-Seerup, K., & Sandelin, A. (2017). The landscape of isoform switches in human cancers. Molecular Cancer Research, 15(9), 1206–1220. https://doi.org/10.1158/1541-7786.MCR-16-0459

Vitting-Seerup, K., & Sandelin, A. (2019). IsoformSwitchAnalyzeR: analysis of changes in genome-wide patterns of alternative splicing and its functional consequences. Bioinformatics (Oxford, England), 35(21), 4469–4471. https://doi.org/10.1093/bioinformatics/btz247

Wan, Y. W., Al-Ouran, R., Mangleburg, C. G., Perumal, T. M., Lee, T. V., Allison, K., Swarup, V., Funk, C. C., Gaiteri, C., Allen, M., Wang, M., Neuner, S. M., Kaczorowski, C. C., Philip, V. M., Howell, G. R., Martini-Stoica, H., Zheng, H., Mei, H., Zhong, X., … Logsdon, B. A. (2020). Meta-Analysis of the Alzheimer’s Disease Human Brain Transcriptome and Functional Dissection in Mouse Models. Cell Reports, 32(2). https://doi.org/10.1016/j.celrep.2020.107908

Wang, M., Beckmann, N. D., Roussos, P., Wang, E., Zhou, X., Wang, Q., Ming, C., Neff, R., Ma, W., Fullard, J. F., Hauberg, M. E., Bendl, J., Peters, M. A., Logsdon, B., Wang, P., Mahajan, M., Mangravite, L. M., Dammer, E. B., Duong, D. M., … Zhang, B. (2018). The Mount Sinai cohort of large-scale genomic, transcriptomic and proteomic data in Alzheimer’s disease. Scientific Data, 5. https://doi.org/10.1038/sdata.2018.185

Xu, J., Patassini, S., Rustogi, N., Riba-Garcia, I., Hale, B. D., Phillips, A. M., Waldvogel, H., Haines, R., Bradbury, P., Stevens, A., Faull, R. L. M., Dowsey, A. W., Cooper, G. J. S., & Unwin, R. D. (2019). Regional protein expression in human Alzheimer’s brain correlates with disease severity. Communications Biology, 2(1). https://doi.org/10.1038/s42003-018-0254-9

Yi, L., Pimentel, H., Bray, N. L., & Pachter, L. (2018). Gene-level differential analysis at transcript-level resolution. Genome Biology, 19(1). https://doi.org/10.1186/s13059-018-1419-z

Yu, N. N., Tan, M. S., Yu, J. T., Xie, A. M., & Tan, L. (2016). The Role of Reelin Signaling in Alzheimer’s Disease. Molecular Neurobiology, 53(8), 5692–5700. https://doi.org/10.1007/s12035-015-9459-9

Zhou, Y., Hayashi, I., Wong, J., Tugusheva, K., Renger, J. J., & Zerbinatti, C. (2014). Intracellular clusterin interacts with brain isoforms of the bridging integrator 1 and with the microtubule-associated protein Tau in Alzheimer’s Disease. PLoS ONE, 9(7). https://doi.org/10.1371/journal.pone.0103187

Zullo, J. M., Drake, D., Aron, L., O’Hern, P., Dhamne, S. C., Davidsohn, N., Mao, C. A., Klein, W. H., Rotenberg, A., Bennett, D. A., Church, G. M., Colaiácovo, M. P., & Yankner, B. A. (2019). Regulation of lifespan by neural excitation and REST. In Nature (Vol. 574, Issue 7778). https://doi.org/10.1038/s41586-019-1647-8

