## Supplementary Material for "Differential transcript usage unravels gene expression alterations in Alzheimer’s disease human brains"

### Supplementary Figures:

**Supplementary Figure 1. Allen Brain Atlas scRNAseq MTG dataset information.** A) tSNE representation showing distribution of single cells in two dimensions; B) Donut plot explaining percentage of cell types among all groups with absolute number of cells; C) Dotplot showing cell type known markers elucidating cell clusters findings; D) Boxplot demonstrating distribution of average expression among all cell types; E) UpsetR exhibiting number of genes expressed in each cell type combination.

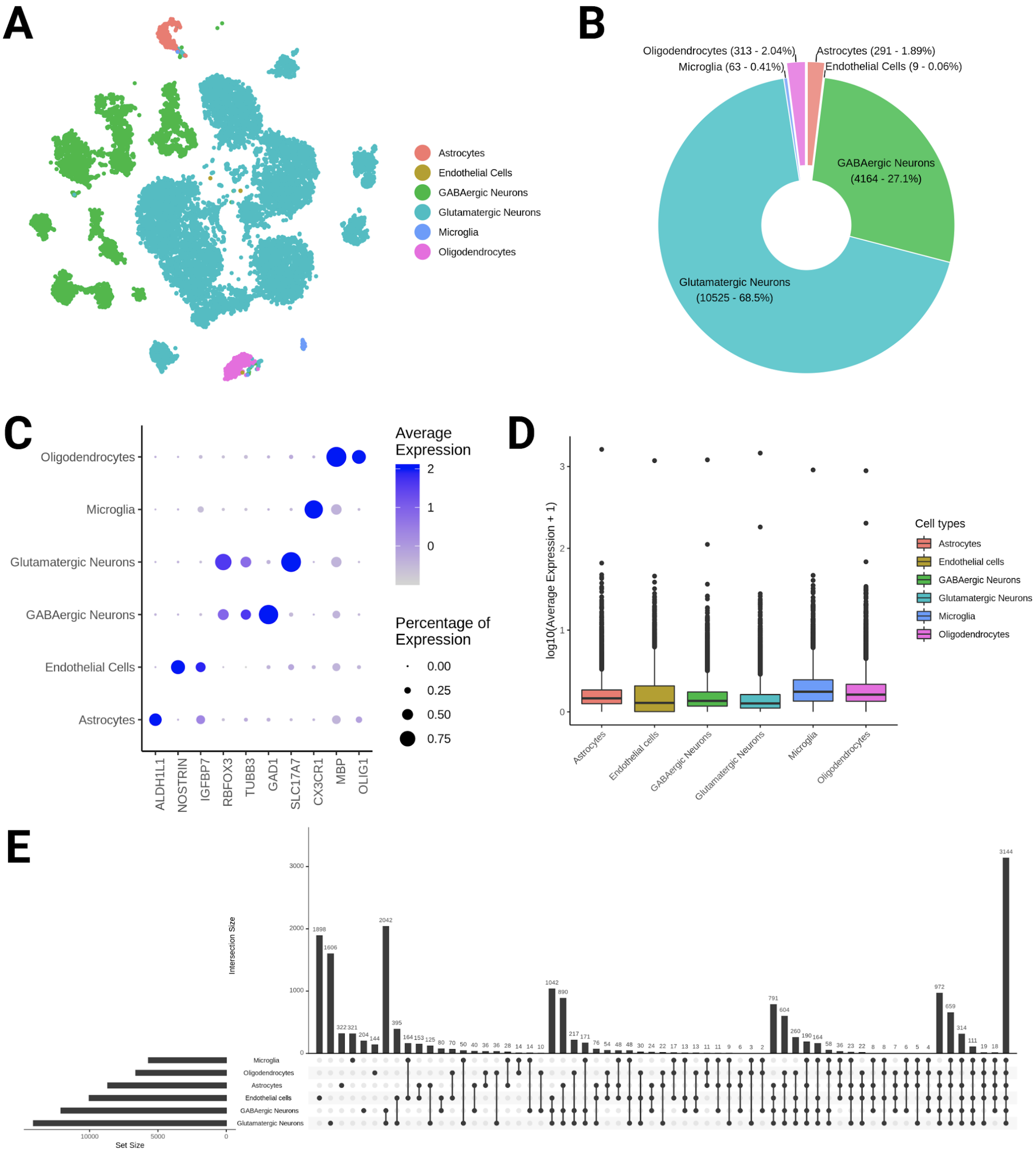
A
Astrocytes
Endothelial Cells
GABAergic Neurons
Glutamatergic Neurons
Microglia
Oligodendrocytes
B
Oligodendrocytes (313 - 2.04%)
Microglia (63 - 0.41%)
Astrocytes (291 - 1.89%)
Endothelial Cells (9 - 0.06%)
GABAergic Neurons (4164 - 27.1%)
Glutamatergic Neurons (10525 - 68.5%)
C
Oligodendrocytes
Microglia
Glutamatergic Neurons
GABAergic Neurons
Endothelial Cells
Astrocytes
ALDH1L1
NOSTRIN
IGFBP7
RBFOX3
TUBB3
GAD1
SLC17A7
CX3CR1
MBP
OLIG1
Average Expression
2
1
0
Percentage of Expression
0.00
0.25
0.50
0.75
D
log10(Average Expression + 1)
Cell types
Astrocytes
Endothelial cells
GABAergic Neurons
Glutamatergic Neurons
Microglia
Oligodendrocytes
E
Intersection Size
Set Size
Microglia
Oligodendrocytes
Astrocytes
Endothelial cells
GABAergic Neurons
Glutamatergic Neurons

**Supplementary Figure 2. scRNAseq signatures comparison with Grubman and Mathys et al works in Glutamatergic Neurons, GABAergic Neurons and Astrocytes.** A,C,E) UpsetR and Venn Diagram illustrate intersection among different studies; B,D,F) Gene-set enrichment analysis (GSEA) with cell-type signatures among all studies. As a threshold for GSEA, FDR < 0.01, intersection size > 3 and precision > 0.05. Analyses were made for glutamatergic neurons (A-B), gabaergic neurons (C-D) and astrocytes (E-F).

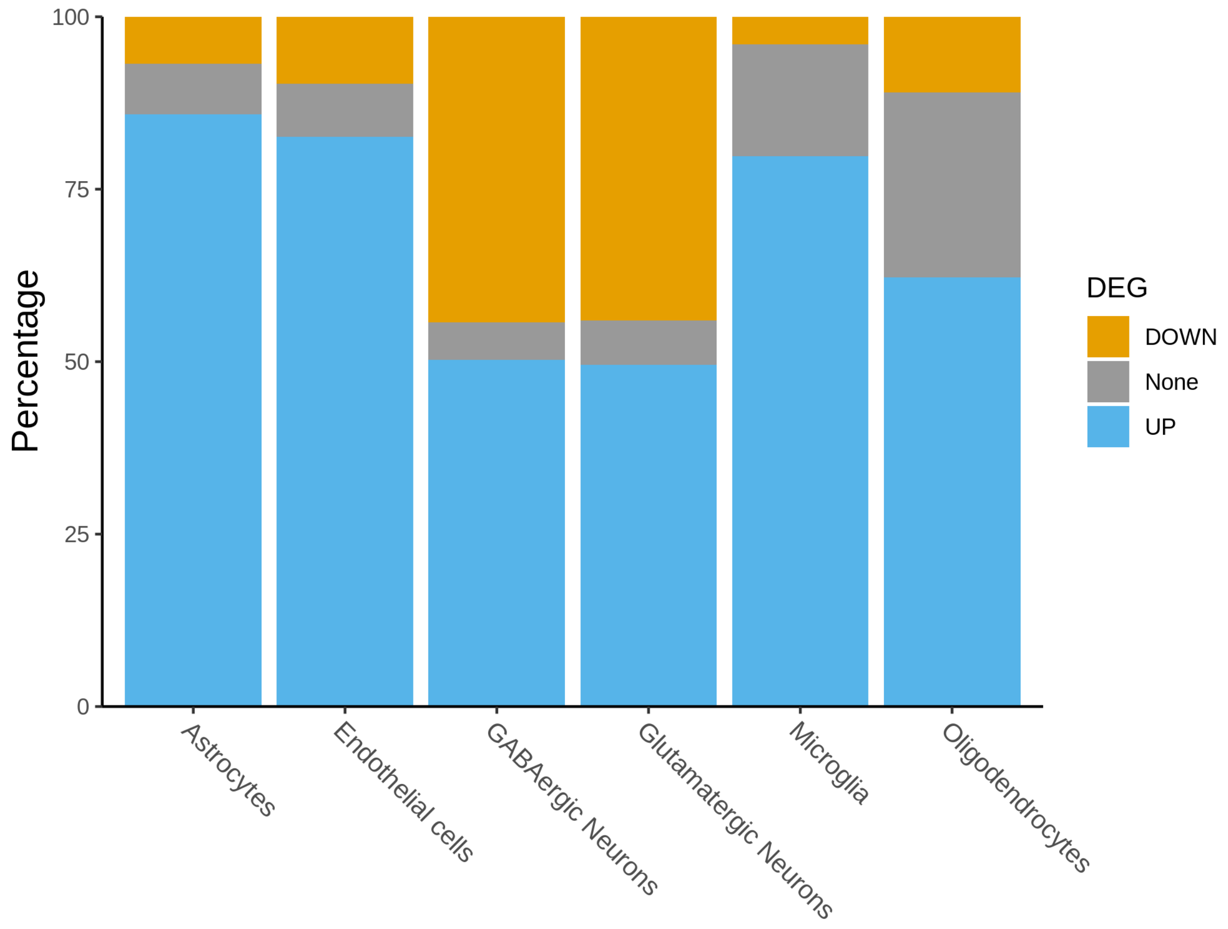

**Supplementary Figure 3. Percentage of DEGs regulation among cell types.** Graphic showing the percentage of genes UP (blue) and DOWN (yellow) regulated in both TL datasets (Mayo and MSBB_TL), as well as genes altered in discordant directions in these two datasets (grey). Observe that neurons have about half of the genes altered in each direction, whereas astrocytes, oligodendrocytes, microglia and endothelial cells have a predominance of up regulated genes.

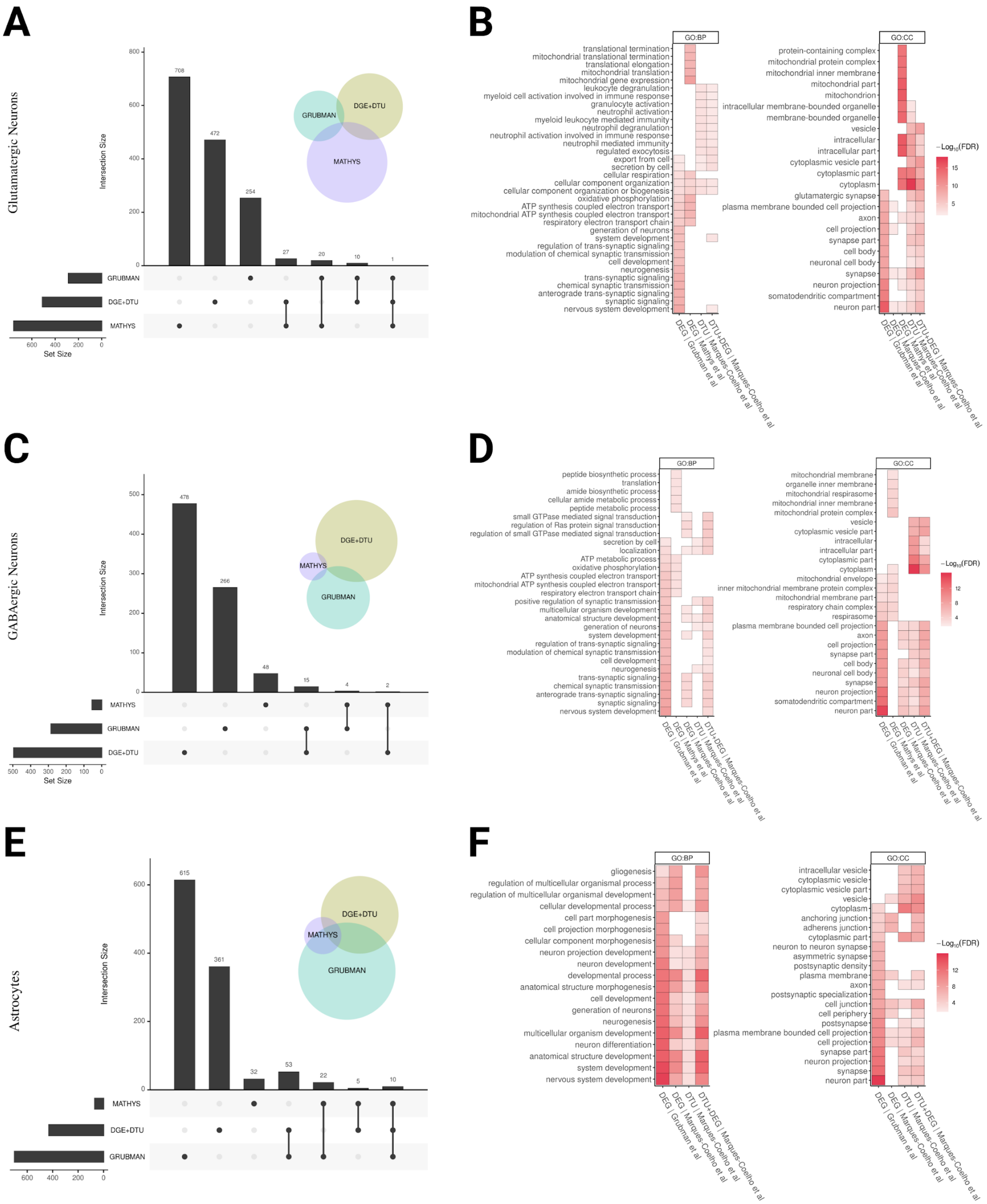

A
Glutamatergic Neurons
Intersection Size
Set Size
GRUBMAN
DGE+DTU
MATHYS
708
472
254
27
20
10
1
B
GO:BP
GO:CC
−Log10(FDR)
translational termination
mitochondrial translational termination
translational elongation
mitochondrial translation
mitochondrial gene expression
leukocyte degranulation
myeloid cell activation involved in immune response
granulocyte activation
neutrophil activation
myeloid leukocyte mediated immunity
neutrophil degranulation
neutrophil activation involved in immune response
neutrophil mediated immunity
regulated exocytosis
export from cell
secretion by cell
cellular respiration
cellular component organization
cellular component organization or biogenesis
oxidative phosphorylation
ATP synthesis coupled electron transport
mitochondrial ATP synthesis coupled electron transport
respiratory electron transport chain
generation of neurons
neurogenesis
system development
regulation of trans-synaptic signaling
modulation of chemical synaptic transmission
cell development
trans-synaptic signaling
chemical synaptic transmission
anterograde trans-synaptic signaling
synaptic signaling
nervous system development
protein-containing complex
mitochondrial protein complex
mitochondrial inner membrane
mitochondrial part
mitochondrion
intracellular membrane-bounded organelle
membrane-bounded organelle
vesicle
intracellular
intracellular part
cytoplasmic vesicle part
cytoplasmic part
cytoplasm
glutamatergic synapse
plasma membrane bounded cell projection
axon
cell projection
synapse part
cell body
neuronal cell body
synapse
neuron projection
somatodendritic compartment
neuron part
DEG | Grubman et al
DEG | Mathys et al
DTU | Marques-Coelho et al
DTU+DEG | Marques-Coelho et al
C
GABAergic Neurons
Intersection Size
Set Size
MATHYS
GRUBMAN
DGE+DTU
478
266
48
15
4
2
D
GO:BP
GO:CC
−Log10(FDR)
peptide biosynthetic process
translation
amide biosynthetic process
cellular amide metabolic process
peptide metabolic process
small GTPase mediated signal transduction
regulation of Ras protein signal transduction
regulation of small GTPase mediated signal transduction
secretion by cell
localization
ATP metabolic process
oxidative phosphorylation
ATP synthesis coupled electron transport
mitochondrial ATP synthesis coupled electron transport
respiratory electron transport chain
positive regulation of synaptic transmission
multicellular organism development
anatomical structure development
generation of neurons
system development
regulation of trans-synaptic signaling
modulation of chemical synaptic transmission
cell development
neurogenesis
trans-synaptic signaling
chemical synaptic transmission
anterograde trans-synaptic signaling
synaptic signaling
nervous system development
mitochondrial membrane
organelle inner membrane
mitochondrial respirasome
mitochondrial inner membrane
mitochondrial protein complex
vesicle
cytoplasmic vesicle part
intracellular
intracellular part
cytoplasmic part
cytoplasm
mitochondrial envelope
inner mitochondrial membrane protein complex
mitochondrial membrane part
respiratory chain complex
respirasome
plasma membrane bounded cell projection
axon
cell projection
synapse part
cell body
neuronal cell body
synapse
neuron projection
somatodendritic compartment
neuron part
DEG | Grubman et al
DEG | Mathys et al
DTU | Marques-Coelho et al
DTU+DEG | Marques-Coelho et al
E
Astrocytes
Intersection Size
MATHYS
DGE+DTU
GRUBMAN
615
361
32
53
22
5
10
F
GO:BP
GO:CC
−Log10(FDR)
gliogenesis
regulation of multicellular organismal process
regulation of multicellular organismal development
cellular developmental process
cell part morphogenesis
cell projection morphogenesis
cellular component morphogenesis
neuron projection development
neuron development
developmental process
anatomical structure morphogenesis
cell development
generation of neurons
neurogenesis
multicellular organism development
neuron differentiation
anatomical structure development
system development
nervous system development
intracellular vesicle
cytoplasmic vesicle
cytoplasmic vesicle part
vesicle
cytoplasm
anchoring junction
adherens junction
cytoplasmic part
neuron to neuron synapse
asymmetric synapse
postsynaptic density
plasma membrane
axon
postsynaptic specialization
cell junction
cell periphery
postsynapse
plasma membrane bounded cell projection
cell projection
synapse part
neuron projection
synapse
neuron part
DEG | Grubman et al
DEG | Mathys et al
DTU | Marques-Coelho et al
DTU+DEG | Marques-Coelho et al

**Supplementary Figure 4. scRNAseq signatures comparison with Grubman and Mathys et al works in Oligodendrocytes, Endothelial cells and Microglia.** A,C,E) Upset plot and Venn Diagram illustrate intersection among different studies; B,D,F) Gene-set enrichment analysis (GSEA) with cell-type signatures among all studies. As a threshold for GSEA, FDR < 0.01, intersection size > 3 and precision > 0.05. Analyses were made for oligodendrocytes (A-B), endothelial cells (C-D) and microglia (E-F).

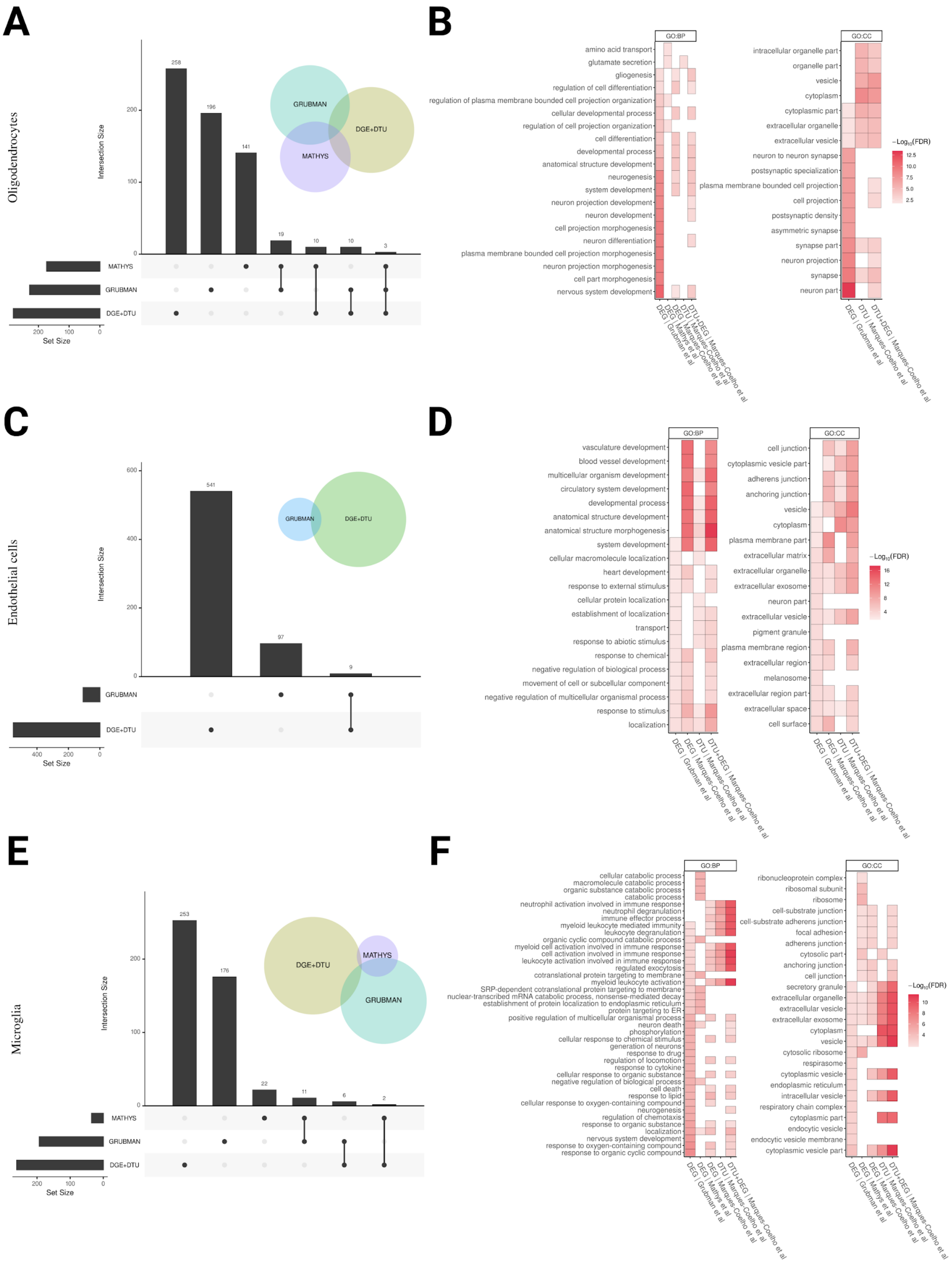

A
Oligodendrocytes
Intersection Size
258
196
141
19
10
10
3
GRUBMAN
DGE+DTU
MATHYS
MATHYS
GRUBMAN
DGE+DTU
Set Size
200
100
0
B
GO:BP
amino acid transport
glutamate secretion
gliogenesis
regulation of cell differentiation
regulation of plasma membrane bounded cell projection organization
cellular developmental process
regulation of cell projection organization
cell differentiation
developmental process
anatomical structure development
neurogenesis
system development
neuron projection development
neuron development
cell projection morphogenesis
neuron differentiation
plasma membrane bounded cell projection morphogenesis
neuron projection morphogenesis
cell part morphogenesis
nervous system development
DEG | Grubman et al
DEG | Mathys et al
DTU | Marques-Coelho et al
DTU+DEG | Marques-Coelho et al
GO:CC
intracellular organelle part
organelle part
vesicle
cytoplasm
cytoplasmic part
extracellular organelle
extracellular vesicle
neuron to neuron synapse
postsynaptic specialization
plasma membrane bounded cell projection
cell projection
postsynaptic density
asymmetric synapse
synapse part
neuron projection
synapse
neuron part
DEG | Grubman et al
DTU | Marques-Coelho et al
DTU+DEG | Marques-Coelho et al
−Log10(FDR)
12.5
10.0
7.5
5.0
2.5
C
Endothelial cells
Intersection Size
600
400
200
0
541
97
9
GRUBMAN
DGE+DTU
GRUBMAN
DGE+DTU
Set Size
400
200
0
D
GO:BP
vasculature development
blood vessel development
multicellular organism development
circulatory system development
developmental process
anatomical structure development
anatomical structure morphogenesis
system development
cellular macromolecule localization
heart development
response to external stimulus
cellular protein localization
establishment of localization
transport
response to abiotic stimulus
response to chemical
negative regulation of biological process
movement of cell or subcellular component
negative regulation of multicellular organismal process
response to stimulus
localization
DEG | Grubman et al
DEG | Marques-Coelho et al
DTU | Marques-Coelho et al
DTU+DEG | Marques-Coelho et al
GO:CC
cell junction
cytoplasmic vesicle part
adherens junction
anchoring junction
vesicle
cytoplasm
plasma membrane part
extracellular matrix
extracellular organelle
extracellular exosome
neuron part
extracellular vesicle
pigment granule
plasma membrane region
extracellular region
melanosome
extracellular region part
extracellular space
cell surface
DEG | Grubman et al
DEG | Marques-Coelho et al
DTU | Marques-Coelho et al
DTU+DEG | Marques-Coelho et al
−Log10(FDR)
16
12
8
4
E
Microglia
Intersection Size
200
100
0
253
176
22
11
6
2
DGE+DTU
MATHYS
GRUBMAN
MATHYS
GRUBMAN
DGE+DTU
Set Size
200
100
0
F
GO:BP
cellular catabolic process
macromolecule catabolic process
organic substance catabolic process
catabolic process
neutrophil activation involved in immune response
neutrophil degranulation
immune effector process
myeloid leukocyte mediated immunity
leukocyte degranulation
organic cyclic compound catabolic process
myeloid cell activation involved in immune response
cell activation involved in immune response
leukocyte activation involved in immune response
regulated exocytosis
cotranslational protein targeting to membrane
myeloid leukocyte activation
SRP-dependent cotranslational protein targeting to membrane
nuclear-transcribed mRNA catabolic process, nonsense-mediated decay
establishment of protein localization to endoplasmic reticulum
protein targeting to ER
positive regulation of multicellular organismal process
neuron death
phosphorylation
cellular response to chemical stimulus
generation of neurons
response to drug
regulation of locomotion
response to cytokine
cellular response to organic substance
negative regulation of biological process
cell death
response to lipid
cellular response to oxygen-containing compound
neurogenesis
regulation of chemotaxis
response to organic substance
localization
nervous system development
response to oxygen-containing compound
response to organic cyclic compound
DEG | Grubman et al
DEG | Mathys et al
DEG | Marques-Coelho et al
DTU | Marques-Coelho et al
DTU+DEG | Marques-Coelho et al
GO:CC
ribonucleoprotein complex
ribosomal subunit
ribosome
cell-substrate junction
cell-substrate adherens junction
focal adhesion
adherens junction
cytosolic part
anchoring junction
cell junction
secretory granule
extracellular organelle
extracellular vesicle
extracellular exosome
cytoplasm
vesicle
cytosolic ribosome
respirasome
cytoplasmic vesicle
endoplasmic reticulum
intracellular vesicle
respiratory chain complex
cytoplasmic part
endocytic vesicle
endocytic vesicle membrane
cytoplasmic vesicle part
DEG | Grubman et al
DEG | Mathys et al
DEG | Marques-Coelho et al
DTU | Marques-Coelho et al
DTU+DEG | Marques-Coelho et al
−Log10(FDR)
10
8
6
4

**Supplementary Figure 5. Expression of BIN1 isoform 1 is reduced in AD brains.** A) Western blot for BIN1 and ACTIN in brain lysates obtained from the frontal cortex (FCx) and hippocampus (Hip) of non-pathology (n=3) and AD (n=6) individuals. Bands corresponding to isoforms 1 and 9 are indicated (Iso1 and Iso9, respectively). B-C) Quantification of total BIN1 normalized per ACTIN (B) and BIN1 isoform 1 normalized by the total BIN1 (C). ANOVA $F_{(3,14)}$= 3.36 and p=0.0494 (B); $F_{(3,14)}$= 16.24 and p<0.0001 (C); Tukey's multiple comparisons test * Padj<0.05 and *** Padj<0.001. D) Western blot for BIN1 in brain lysates from 3 control and 6 AD individuals (Hip and FCx samples are intercalated) and HEK cells transfected with lentiviral vector carrying plasmids encoding for BIN1 isoforms 1 (Iso1) and 9 (Iso9). Observe the correspondence between the first and third band in the brain (lanes 1 and 2) with isoform 1 (lane 3) and 9 (lane 4), respectively.

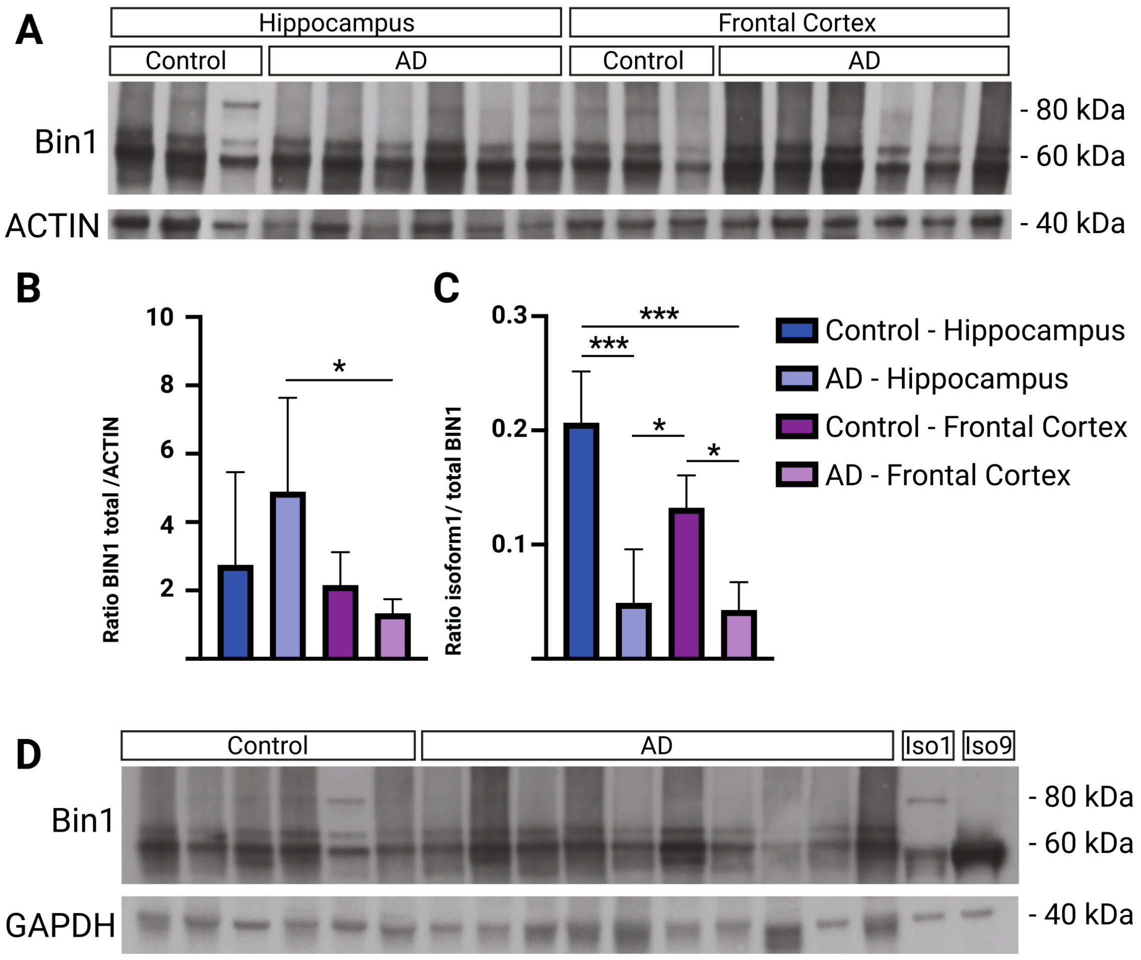

### Description of supplementary tables:

**Supplementary table 1.** Summary statistics for DEG and DTU analyses

**Supplementary table 2.** List of DEGs and gDTUs in TLI and FLI

**Supplementary table 3.** GSEA for DEGs and gDTUs in the temporal lobe intersection (TLI)

**Supplementary table 4.** Summary of splicing events (SE) and events consequences (EC) observed in different datasets

**Supplementary table 5.** Summary statistics for DTU analysis in MSBB using Braak stages

**Supplementary table 6.** List of splicing related genes used in this study and their corresponding ontology terms

**Supplementary table 7.** Expression of genes identified in DEGs and DTU analyses in different cell types/subtypes

**Supplementary table 8.** GSEA for DEGs and gDTUs in the TLI per cell type

**Supplementary table 9.** GSEA for DEGs identified in different cell types in previous single-cell RNAseq studies

**Supplementary table 10.** List of AD risk genes used in this study

**Supplementary table 11.** Expression of AD risk/causal factors in different cell types/subtypes

S**upplementary table 12.** Summary of clinico-pathological criteria used in different consortia to classify AD and control subjects

**Supplementary table 13.** Metadata for all individual samples used in this study
